# Leukemogenesis in Acute Lymphoblastic Leukemia through the Lens of Developmental Dynamics of Lymphoid Progenitor Cells: a systematic review

**DOI:** 10.1101/2024.07.14.24310322

**Authors:** Ovais Shafi, Rahimeen Rajpar, Finza Kanwal, Muhammad Waqas, Osama Jawed Khan, Raveena, Ankash Kumar, Abdul Jabbar, Manwar Madhwani

## Abstract

**Objective:** The objective of this study is to investigate how lymphoid progenitor mechanisms contribute to the leukemogenesis in Acute Lymphoblastic Leukemia.

**Background:** Acute Lymphoblastic Leukemia (ALL), the leading childhood cancer, remains challenging despite treatment advances. ALL originates from aberrant clonal expansion of immature lymphoid progenitor cells (LPCs) due to genetic abnormalities. This study looks into LPC genetic dysregulations, aiming to identify therapeutic targets and enhance prognostic stratification. Understanding ALL pathogenesis not only improves outcomes but also informs broader cancer research, offering potential insights for hematologic malignancies and oncogenesis, promising advancements in clinical practice.

**Methods:** Databases, including PubMed, MEDLINE, Google Scholar, and open access/subscription-based journals were searched for published articles without any date restrictions, to investigate leukemogenesis in acute lymphoblastic leukemia and the impact of down syndrome through the developmental regulators of lymphoid progenitor cells. Based on the criteria mentioned in the methods section, studies were systematically reviewed to investigate ALL leukemogenesis. This study adheres to relevant PRISMA guidelines (Preferred Reporting Items for Systematic Reviews and Meta-Analyses).

**Results:** Acute Lymphoblastic Leukemia (ALL) development is marked by dysregulations in key factors governing Lymphoid Progenitor Cells (LPCs) proliferation and differentiation. Dysregulations include upregulation of transcription factors Ikaros and PU.1, along with heightened expression of surface markers CD34 and CD38. Increased levels of signaling molecules IL-7, SCF, and FLT3 Ligand drive LPC proliferation, while alterations in epigenetic modifiers DNMTs, HDACs, and HATs further contribute to uncontrolled cell division. Dysregulated transcription factors Ikaros, PU.1, E2A (TCF3), EBF1, and Notch1 disrupt LPC differentiation pathways, alongside aberrant expression of surface markers CD10, CD7, and CD45RA. Dysregulated signaling through IL-7 and the Notch pathway, along with epigenetic modifications mediated by DNMTs, HDACs, and HATs, collectively create a conducive landscape for ALL development.

**Conclusion:** This study into the origins of ALL investigates the roles of transcription factors, surface markers, signaling molecules, and epigenetic modifiers in LPC proliferation and differentiation. Dysregulation of these components, often due to mutations in genes like GATA1, CRLF2, JAK2, and RAS, disrupts normal hematopoietic development and leads to leukemic transformation.

## Background

Acute Lymphoblastic Leukemia (ALL) is the most common childhood cancer, accounting for approximately 25% of all pediatric malignancies [1]. Despite significant advances in treatment, ALL remains a challenging disease to manage, with a subset of patients experiencing relapse or refractory disease. Understanding the underlying mechanisms driving ALL development is crucial for improving patient outcomes and developing more effective therapeutic strategies [2]. ALL arises from the clonal expansion of immature lymphoid progenitor cells (LPCs) in the bone marrow, leading to the accumulation of abnormal lymphoblasts [3]. While the exact etiology of ALL is multifactorial and heterogeneous, emerging evidence suggests that genetic abnormalities play a central role in disease pathogenesis. Dysregulations in the genetic architecture of LPCs, including mutations in transcription factors, surface markers, signaling molecules, and epigenetic modifiers, can disrupt normal hematopoietic development and predispose LPCs to leukemic transformation [4].

This study aims to investigate the complex interplay between genetic alterations and ALL development by dissecting the molecular pathways involved in LPC proliferation and differentiation [5]. By characterizing the aberrant genetic signatures associated with ALL, we can identify new therapeutic targets and develop targeted therapies tailored to the specific molecular abnormalities present in individual patients [6]. Additionally, understanding the impact of genetic dysregulations on LPC behavior can inform prognostic stratification and guide treatment decisions, ultimately improving patient outcomes. Furthermore, this research has implications beyond ALL, with potential implications for other hematologic malignancies and cancer types [7]. By focusing on the genetic underpinnings of ALL, we can gain insights into the broader mechanisms of oncogenesis and identify common molecular targets for therapeutic intervention. Thus, this study holds promise for advancing our understanding of oncogenesis and translating discoveries into clinical practice to benefit patients worldwide. Findings emerging through research in this area of ALL have translational potential, with implications for the broader field of oncology. Insights gained from studying the genetic and molecular underpinnings of ALL can inform research and clinical practice across various cancer types, leading to advancements in diagnosis, treatment, and patient care [8].

Down Syndrome (DS), a genetic disorder resulting from the triplication of chromosome 21, is associated with a significantly increased risk of developing Acute Lymphoblastic Leukemia (ALL). While ALL is the most common childhood cancer, individuals with DS exhibit a 10 to 20-fold higher incidence of this malignancy compared to the general population [9]. The underlying mechanisms linking DS to ALL susceptibility are complex and multifactorial. Several genetic alterations associated with DS, including mutations in genes such as GATA1 and alterations in the CRLF2 gene, have been implicated in ALL leukemogenesis [10]. Additionally, associated mutations affecting pathways involving either the JAK2 or RAS genes further contribute to the heightened risk of ALL in individuals with DS. These genetic abnormalities disrupt the normal development and function of lymphoid progenitor cells (LPCs), leading to their malignant transformation and the development of ALL. Understanding the molecular pathways linking DS to ALL susceptibility is critical for elucidating the underlying mechanisms of leukemogenesis in this population. Furthermore, it has significant implications for both clinical management and therapeutic interventions [11]. By focusing on the critical interplay between genetic abnormalities and ALL development in individuals with DS, transformative therapeutic targets can be identified, leading to improved treatment strategies and better outcomes for this high-risk patient population. Therefore, investigating the role of DS in increasing the risk of ALL provides valuable insights into the pathogenesis of this disease and offers opportunities for personalized approaches to its management [12, 13].

## Methods

### Aim of the Study

The aim of this study is to look into the roles of specific transcription factors, surface markers, and signaling pathways in the development of lymphoid progenitor cells and their implications in the leukemogenesis of Acute Lymphoblastic Leukemia (ALL). This includes examining the impact of dysregulations in these factors on the disruption of cell fate, the mechanisms leading to ALL, and the influence of Down Syndrome (DS)-associated mutations on ALL leukemogenesis and risk. These are also the limitations of the study.

### Research Question

How do key transcription factors, surface markers, and signaling pathways of lymphoid progenitor cells contribute to the leukemogenesis of Acute Lymphoblastic Leukemia (ALL), also in the context of Down Syndrome?

### Search Focus

A comprehensive literature search was conducted using the PUBMED database, MEDLINE database, and Google Scholar, as well as open access and subscription-based journals. There were no date restrictions for published articles.

The search focused on the following key transcription factors, surface markers, and signaling pathways involved in the development of lymphoid progenitor cells, specifically investigating their roles in Acute Lymphoblastic Leukemia (ALL) leukemogenesis, including in the context of Down Syndrome (DS) in ALL.

#### Transcription Factors and Surface Markers

Ikaros, PU.1, E2A (TCF3), EBF1, Notch1, CD34, CD38, CD10, CD7, CD45RA, FLT3 (CD135)

#### Signaling Pathways and Factors

Interleukin-7, Stem Cell Factor, Epigenetic Modifiers.

Screening of the literature was also done on this same basis and related data was extracted. Literature search began in August 2020 and ended in November 2023. An in-depth investigation was conducted during this duration based on the parameters of the study as defined above. During revision, further literature was searched and referenced until May 2024. The literature search and all sections of the manuscript were checked multiple times during the months of revision (December 2023 – May 2024) to maintain the highest accuracy possible. This comprehensive approach ensured that the selected studies provided valuable insights into the developmental biology of lymphoid progenitor cells and their implications in ALL leukemogenesis, including DS-related mutations. This study adheres to relevant PRISMA guidelines (Preferred Reporting Items for Systematic Reviews and Meta-Analyses).

### Search Queries/Keywords

1. General Terms:

“Acute Lymphoblastic Leukemia” OR “ALL“
“ALL leukemogenesis“
“Lymphoid Progenitor Cells” OR “LPCs“
“Down Syndrome” AND “ALL“
2. Transcription Factors:

“Ikaros” AND “ALL“
“PU.1” AND “ALL leukemogenesis“
“E2A” OR “TCF3” AND “ALL“
“EBF1” AND “ALL“
3. Surface Markers:

“CD34” AND “ALL“
“CD38” AND “ALL leukemogenesis“
“CD10” AND “ALL“
“CD7” AND “lymphoid progenitor cells“
“CD45RA” AND “leukemia“
“FLT3” OR “CD135” AND “ALL“
4. Signaling Pathways and Factors:

“Interleukin-7” OR “IL-7” AND “ALL“
“Stem Cell Factor” AND “ALL“
“Epigenetic modifiers” AND “ALL“
“Notch signaling pathway” AND “ALL“
5. Down Syndrome Specific Searches:

“Down Syndrome” AND “leukemia“
“Impact of Down Syndrome” AND “ALL“
“GATA1 mutations” AND “Down Syndrome” AND “ALL“
“CRLF2” AND “Down Syndrome” AND “ALL“
“JAK2 mutations” AND “Down Syndrome” AND “ALL“
“RAS mutations” AND “Down Syndrome” AND “ALL“

Boolean operators (AND, OR) were utilized to construct search queries in relation to the following terms: “Acute Lymphoblastic Leukemia” (ALL), “Lymphoid Progenitor Cells” (LPC), “Down Syndrome,” and “Leukemogenesis.” These operators were applied in relation to key transcription factors, surface markers, and signaling pathways involved in the developmental dynamics of lymphoid progenitor cells. This approach facilitated the comprehensive retrieval of relevant articles by combining these terms to capture studies focusing on their interrelations and individual contributions to leukemogenesis.

### Objectives of the Searches

- To determine the impact of dysregulations in disrupting the cell fate of lymphoid progenitor cells.
- To look into the damage leading to ALL leukemogenesis.
- To assess Down Syndrome based mutations (GATA1, CRLF2, JAK2, RAS) in ALL leukemogenesis and their role in increasing ALL risk.

### Screening and Eligibility Criteria

#### Initial Screening

Articles were initially screened based on titles and abstracts to identify direct relevance to the study objectives.

#### Full-Text Review

Articles that passed the initial screening underwent a comprehensive full-text review. Articles were included if they provided valuable insights into the roles of the specified transcription factors, surface markers, and signaling pathways in ALL leukemogenesis and DS-related mutations in ALL.

#### Data Extraction

Relevant data was extracted from each selected article, focusing on key findings and outcomes related to the study objectives.

##### Inclusion and Exclusion Criteria

###### Inclusion Criteria

Articles directly related to the key transcription factors, surface markers, and signaling pathways involved in the development of lymphoid progenitor cells. Studies focusing on the impact of dysregulations on cell fate, ALL leukemogenesis, and DS-related mutations in ALL.

###### Exclusion Criteria

Articles that did not conform to the study focus. Insufficient methodological rigor. Data not aligning with the research questions.

### Rationale for Screening and Inclusion

- **Ikaros:** Essential for lymphoid lineage commitment and differentiation. Regulates development of B and T cells. Loss or mutation leads to impaired lymphoid differentiation and contributes to leukemogenesis.
- **PU.1:** Critical for development of myeloid and lymphoid lineages. Influences early B-cell development. Altered expression disrupts normal hematopoiesis, promoting leukemic cell proliferation and survival.
- **E2A (TCF3):** Crucial for B-cell lineage specification and differentiation. Mutations or translocations lead to aberrant cell growth and differentiation.
- **EBF1:** Essential for B-cell commitment and differentiation. Works in concert with E2A. Dysregulation disrupts normal B-cell development, facilitating ALL progression.
- **Notch1:** Regulates T-cell development and differentiation. Mutations frequently observed in T-cell ALL (T-ALL), leading to uncontrolled cell proliferation and survival.
- **CD34:** Marker of hematopoietic stem and progenitor cells, including lymphoid progenitors. High expression associated with leukemic stem cells, indicating a role in maintaining the leukemic clone.
- **CD38:** Marker of early lymphoid and myeloid progenitors. Involved in cell adhesion and signaling. Aberrant expression contributes to leukemic cell proliferation and survival.
- **CD10:** Marker of early B-cell progenitors. High expression characteristic of precursor B-ALL, aiding in leukemic cell identification and classification.
- **CD7:** Early marker of T-cell progenitors. Overexpression common in T-ALL, indicating involvement in leukemic cell proliferation.
- **CD45RA:** Isoform of CD45, involved in lymphocyte signaling. Aberrant expression affects leukocyte signaling pathways, contributing to ALL pathogenesis.
- **FLT3 (CD135):** Receptor tyrosine kinase important for hematopoietic progenitor cell development. Mutations associated with poor prognosis in ALL, leading to increased proliferation and survival of leukemic cells.
- **Interleukin-7 (IL-7):** Crucial for survival and proliferation of early lymphoid progenitors. Dysregulated signaling promotes survival and proliferation of leukemic cells in ALL.
- **Stem Cell Factor (SCF):** Important for hematopoietic stem cell maintenance and differentiation. Abnormal signaling contributes to maintenance and expansion of leukemic stem cells.
- **Epigenetic Modifiers:** Regulate gene expression through DNA methylation and histone modification. Epigenetic dysregulation common in ALL, leading to altered gene expression and leukemogenesis.
- **Down Syndrome-Related Mutations (GATA1, CRLF2, JAK2, RAS):** Frequently observed in ALL patients with Down Syndrome. Contribute to an increased risk of leukemogenesis.

This comprehensive screening and inclusion rationale ensure that the selected studies provide valuable insights into the roles of these critical factors in the development of lymphoid progenitor cells and their implications in ALL leukemogenesis, with a particular focus on Down Syndrome-associated mutations.

### Exclusion Criteria

To ensure that only the most relevant and high-quality studies were included in this investigation of the roles of specific transcription factors, surface markers, and signaling pathways in lymphoid progenitor cell development and ALL leukemogenesis, the following exclusion criteria were applied:

#### 1. Non-Conformity with Study Focus

Articles that did not specifically investigate the roles of Ikaros, PU.1, E2A (TCF3), EBF1, Notch1, CD34, CD38, CD10, CD7, CD45RA, FLT3 (CD135), Interleukin-7, Stem Cell Factor, and Epigenetic Modifiers in lymphoid progenitor cells and ALL leukemogenesis. Studies that did not address the impact of dysregulations in these factors on the cell fate of lymphoid progenitor cells or their roles in ALL leukemogenesis.

#### 2. Insufficient Methodological Rigor

Studies with poor experimental design, lack of appropriate controls, or inadequate statistical analysis. Articles that did not provide clear and reproducible results.

#### 3. Lack of Relevance to Down Syndrome in ALL

Studies that did not explore the involvement of DS-related mutations (GATA1, CRLF2, JAK2, RAS) in ALL leukemogenesis or their role in increasing the risk of ALL in Down Syndrome patients.

#### 4. Incomplete or Irrelevant Data

Articles that did not provide sufficient data or detailed analysis related to the key transcription factors, surface markers, and signaling pathways under investigation. Studies that focused on unrelated biological processes or diseases.

#### 5. Duplicate Publications

Duplicate articles or those that presented redundant data from previously included studies.

#### 6. Non-English Publications

Articles not published in the English language, to ensure consistency and comprehensibility of the reviewed literature.

#### 7. Unpublished Studies

Unpublished studies, including conference abstracts and dissertations were not included, to ensure the inclusion of rigorously validated research.

#### 8. Date of Publication

Although there were no date restrictions for the literature search, studies that did not align with current understanding and technological advancements were critically assessed for their relevance and methodological robustness.

By applying these exclusion criteria, the study maintained a high standard of evidence, ensuring that the included literature provided valuable and relevant insights into the roles of the specified transcription factors, surface markers, and signaling pathways in lymphoid progenitor cell development and ALL leukemogenesis, with a particular focus on Down Syndrome-associated mutations.

### Assessment of Article Quality and Potential Biases

Ensuring the quality and minimizing potential biases of the selected articles were crucial aspects to guarantee the rigor and reliability of the research findings.

- **Quality Assessment:** The initial step in quality assessment involved evaluating the methodological rigor of the selected articles. This included a thorough examination of the study design, data collection methods, and analyses conducted. The significance of the study’s findings was weighed based on the quality of the evidence presented. Articles demonstrating sound methodology—such as well-designed studies, controlled variables, and scientifically robust data—were considered of higher quality. Peer-reviewed articles, scrutinized by experts in the field, served as a significant indicator of quality.
- **Potential Biases Assessment:**

○ **Publication Bias:** To address the potential for publication bias, a comprehensive search strategy was adopted to include a balanced representation of both positive and negative results, incorporating a wide range of published articles from databases like Google Scholar.
○ **Selection Bias:** Predefined and transparent inclusion criteria were applied to minimize subjectivity in the selection process. Articles were chosen based on their relevance to the study’s objectives, adhering strictly to these criteria. This approach reduced the risk of subjectivity and ensured that the selection process was objective and consistent.
○ **Reporting Bias:** To mitigate reporting bias, articles were checked for inconsistencies or missing data. Multiple detailed reviews of the methodologies and results were conducted for all selected articles to identify and address any reporting bias.

By including high-quality, peer-reviewed studies and thoroughly assessing potential biases, this study aimed to provide a robust foundation for the results and conclusions presented.

### Language and publication restrictions

We restricted our selection to publications in the English language. There were no limitations imposed on the date of publication. Unpublished studies were not included in our analysis.

## Results

A total of 3483 articles were identified using database searching, and 3322 were recorded after duplicates removal. 2944 were excluded after screening of title/abstract, 102 were finally excluded, and 3 articles were excluded during data extraction. These exclusions were primarily due to factors such as non-conformity with the study focus, insufficient methodological rigor, or data that did not align with our research questions. Finally, 273 articles were included as references.

### Investigating ALL Leukemogenesis Through the Lens of Lymphoid Progenitor Cells

#### 1. Ikaros Transcription Factor

##### Impact of Dysregulations in Disrupting the Cell Fate of Lymphoid Progenitor Cells

Dysregulation of Ikaros, a critical transcription factor involved in the developmental biology of Lymphoid Progenitor Cells (CLPs), can significantly impact the cell fate of these progenitors, potentially leading to the development of Acute Lymphoblastic Leukemia (ALL) [14]. One mechanism by which dysregulation in Ikaros may disrupt the cell fate of Lymphoid Progenitor Cells towards ALL is aberrant lineage commitment. Ikaros plays a key role in regulating lineage commitment by promoting the differentiation of CLPs into mature lymphoid cells while suppressing alternative lineage fates, such as myeloid differentiation. Dysregulation of Ikaros may disrupt this balance, leading to aberrant lineage commitment and the expansion of progenitors with leukemogenic potential. Another mechanism is impaired differentiation [15]. Proper differentiation of CLPs into mature lymphoid cells is essential for maintaining normal hematopoiesis. Ikaros regulates the expression of genes involved in lymphoid differentiation, ensuring the progression from progenitors to mature lymphoid cells [16]. Dysregulated Ikaros activity may impair this differentiation process, resulting in the accumulation of immature or undifferentiated cells characteristic of ALL. Additionally, Ikaros acts as a tumor suppressor by inhibiting cell proliferation and promoting apoptosis in response to cellular stress [17]. Dysregulation of Ikaros, such as mutations or deletions, may confer a proliferative advantage to CLPs, allowing for uncontrolled cell growth and the expansion of leukemic clones, a hallmark of ALL. Ikaros also plays a critical role in maintaining genomic stability by regulating DNA repair mechanisms and chromatin remodeling. Dysregulated Ikaros function may lead to genomic instability, resulting in the accumulation of genetic mutations and chromosomal abnormalities in CLPs, which can drive leukemogenesis in ALL [18]. Moreover, Ikaros regulates apoptosis by modulating the expression of pro- and anti-apoptotic genes, ensuring the elimination of damaged or aberrant cells. Dysregulation of Ikaros may confer resistance to apoptosis in CLPs, allowing leukemic cells to evade cell death signals and persist, contributing to the progression of ALL [19, 20].

##### Damage leading to ALL Leukemogenesis

Understanding how dysregulated Ikaros alters downstream genes, transcription factors (TFs), and signaling pathways towards a pathologic state is crucial for elucidating the mechanisms underlying the development of Acute Lymphoblastic Leukemia (ALL) leukemogenesis [21]. Damaged Ikaros can significantly impact these aspects. Normally, Ikaros acts as a transcriptional regulator, binding to specific DNA sequences and controlling the expression of target genes involved in lymphoid development and differentiation [22]. When Ikaros is dysregulated or its function is lost, there can be aberrant expression of downstream genes critical for lymphoid differentiation and leukemogenesis. This may involve the downregulation of genes that promote lymphoid differentiation or the upregulation of genes associated with cell proliferation and survival, thereby promoting leukemogenesis. In its normal role, Ikaros interacts with other transcription factors to regulate gene expression and cell fate decisions during lymphoid development. Dysregulated Ikaros disrupts these interactions, leading to altered transcription factor activity [23]. For example, impaired Ikaros function may result in the dysregulation of transcription factors involved in lineage commitment and differentiation, such as EBF1, E2A, and NOTCH1, which contributes to the pathogenesis of ALL. Ikaros also integrates signals from various signaling pathways, including Notch, Wnt, and cytokine signaling, to regulate lymphoid development and differentiation. When Ikaros is dysregulated, it perturbs these signaling pathway activities, leading to aberrant cellular responses. For instance, impaired Ikaros function may dysregulate Notch signaling, a critical pathway in lymphoid development, resulting in enhanced cell proliferation and survival, impaired differentiation, and consequently, the promotion of ALL development. Additionally, Ikaros regulates chromatin accessibility and histone modifications, influencing gene expression patterns during lymphoid development [24]. Dysregulated Ikaros can alter the epigenetic landscape of CLPs, leading to aberrant gene expression profiles that are conducive to leukemogenesis. For example, impaired Ikaros function may result in altered histone modifications or DNA methylation patterns, which in turn dysregulates the expression of genes involved in cell cycle control, apoptosis, and self-renewal. Damaged Ikaros disrupts downstream genes, transcription factors, and signaling pathways involved in lymphoid development, differentiation, and leukemogenesis [25]. Understanding the specific molecular mechanisms by which dysregulated Ikaros contributes to ALL pathogenesis is essential for identifying potential therapeutic targets and developing targeted therapies for the treatment of ALL [26].

##### Down Syndrome based mutations (GATA1, CRLF2, JAK2, RAS) in ALL Leukemogenesis and in increasing ALL Risk

Down Syndrome (DS) is associated with an increased risk of developing Acute Lymphoblastic Leukemia (ALL), and mutations in genes such as GATA1, CRLF2, JAK2, and RAS have been implicated in DS-ALL leukemogenesis [27]. Investigating how these mutations contribute to the dysregulation of Ikaros expression in Lymphoid Progenitor Cells (CLPs) and ALL leukemogenesis involves understanding their interplay with Ikaros and the impact of Down Syndrome on Ikaros expression. Down Syndrome is characterized by trisomy of chromosome 21, which harbors the IKZF1 gene encoding Ikaros [28]. Increased gene dosage due to trisomy 21 may lead to overexpression of Ikaros in CLPs, potentially altering its regulatory functions and downstream effects on lymphoid development. GATA1 mutations in DS-ALL have been associated with downregulation of Ikaros expression [29]. GATA1 directly regulates Ikaros expression, and mutations in GATA1 can disrupt its regulatory function, leading to decreased Ikaros levels. Reduced Ikaros expression due to GATA1 mutations, compounded by potential dosage effects in Down Syndrome, may disrupt normal lymphoid development and differentiation, promoting leukemic transformation in CLPs and contributing to ALL leukemogenesis [30]. Dysregulated CRLF2 signaling in DS-ALL can impact Ikaros expression indirectly by altering signaling pathways involved in its regulation. High CRLF2 levels have been associated with decreased Ikaros expression. Altered CRLF2 signaling and reduced Ikaros expression, in the context of increased gene dosage in Down Syndrome, may further perturb normal lymphoid development, leading to the expansion of leukemic progenitors with enhanced proliferation and reduced differentiation, thereby contributing to ALL leukemogenesis. JAK2 mutations in DS-ALL activate downstream signaling pathways, including the JAK/STAT pathway, which can modulate Ikaros expression indirectly [31]. Dysregulated JAK2 signaling has been associated with decreased Ikaros expression levels. Aberrant JAK2 signaling and reduced Ikaros expression, in the context of increased gene dosage in Down Syndrome, may further promote leukemic transformation by altering the balance between proliferation and differentiation in CLPs, leading to ALL leukemogenesis. Mutations in the RAS pathway, such as activating mutations in RAS genes, can influence Ikaros expression through downstream signaling cascades involved in its regulation. Dysregulated RAS signaling has been associated with decreased Ikaros expression [32]. Dysregulated RAS signaling and reduced Ikaros expression, exacerbated by increased gene dosage in Down Syndrome, may disrupt normal lymphoid development, leading to the expansion of leukemic progenitors with enhanced proliferation, survival, and genomic instability, thereby contributing to ALL leukemogenesis. Mutations in GATA1, CRLF2, JAK2, and RAS associated with Down Syndrome-Associated Acute Lymphoblastic Leukemia (DS-ALL) can contribute to the dysregulation of Ikaros expression in CLPs and ALL leukemogenesis [33]. The interplay between these mutations and the increased gene dosage of Ikaros in Down Syndrome may further exacerbate the disruption of normal lymphoid development, leading to the development of ALL [34].

#### 2. PU.1 Transcription Factor

##### Impact of Dysregulations in Disrupting the Cell Fate of Lymphoid Progenitor Cells

PU.1 is a critical transcription factor involved in the developmental biology of Lymphoid Progenitor Cells (CLPs), playing a critical role in regulating lineage commitment, differentiation, and cell fate decisions. Dysregulations in PU.1 expression or function can significantly impact the cell fate of CLPs, potentially leading to the development of Acute Lymphoblastic Leukemia (ALL) [35]. PU.1 is involved in lineage commitment by promoting the differentiation of CLPs into myeloid and lymphoid lineages. It acts as a key regulator of myeloid differentiation while inhibiting lymphoid lineage commitment. Dysregulated PU.1 expression or function may lead to aberrant lineage commitment, skewing the balance towards myeloid differentiation over lymphoid lineage commitment [36]. This imbalance can result in the expansion of myeloid progenitors at the expense of lymphoid progenitors, predisposing to ALL development. PU.1 regulates the expression of genes essential for lymphoid differentiation, ensuring the proper maturation of CLPs into mature lymphoid cells. Dysregulated PU.1 activity may impair lymphoid differentiation, leading to the accumulation of immature or undifferentiated lymphoid progenitors [37]. This block in differentiation can create a pool of precursor cells with self-renewal capabilities, contributing to the leukemic transformation observed in ALL. PU.1 acts as a regulator of cell proliferation, maintaining the balance between proliferation and differentiation in hematopoietic progenitor cells [38]. Dysregulated PU.1 expression or function may confer a proliferative advantage to CLPs, promoting uncontrolled cell proliferation and the expansion of leukemic clones. This dysregulated proliferation can lead to the accumulation of leukemic blasts characteristic of ALL. PU.1 regulates the expression of genes involved in DNA repair and maintenance of genomic stability [39]. Dysregulated PU.1 activity may lead to genomic instability, resulting in the accumulation of genetic mutations and chromosomal abnormalities in CLPs. This genomic instability can drive leukemogenesis by promoting the acquisition of additional mutations necessary for ALL development. PU.1 governs cell fate decisions in hematopoietic progenitors, directing them towards myeloid or lymphoid lineages. Dysregulated PU.1 expression or function may disrupt normal cell fate decisions, leading to the emergence of leukemic progenitors with altered differentiation potential. This altered cell fate decision-making can contribute to the development of ALL by promoting the expansion of leukemic clones with self-renewal capabilities. Dysregulations in PU.1 expression or function can disrupt normal lymphoid development and predispose Lymphoid Progenitor Cells to ALL development [40].

##### Damage leading to ALL Leukemogenesis

Dysregulation of PU.1 can lead to the development of Acute Lymphoblastic Leukemia (ALL) by altering downstream genes, transcription factors (TFs), and signaling pathways critical for normal lymphoid development [41]. PU.1 regulates the expression of genes involved in myeloid and lymphoid lineage commitment, differentiation, and cell fate decisions. Dysregulated PU.1 activity may result in aberrant expression of downstream genes crucial for lymphoid development [42]. This dysregulation can lead to the upregulation of genes promoting myeloid differentiation and proliferation, while suppressing genes involved in lymphoid differentiation, thereby favoring the development of ALL. PU.1 interacts with various TFs to regulate gene expression and cell fate decisions during hematopoiesis [43]. Dysregulated PU.1 may disrupt interactions with its binding partners, altering TF activity. For example, impaired PU.1 function may lead to dysregulated activity of TFs involved in myeloid and lymphoid lineage specification, contributing to the dysregulated gene expression patterns observed in ALL. PU.1 integrates signals from various signaling pathways, such as cytokine and growth factor signaling, to regulate hematopoietic progenitor cell fate decisions. Dysregulated PU.1 can perturb signaling pathway activity, leading to aberrant cellular responses [44]. For instance, impaired PU.1 function may dysregulate Notch, Wnt, or cytokine signaling pathways critical for lymphoid development, promoting abnormal proliferation and differentiation of progenitor cells and contributing to ALL pathogenesis. PU.1 influences chromatin accessibility and histone modifications, thereby modulating gene expression during hematopoiesis. Dysregulated PU.1 activity may lead to alterations in chromatin structure and epigenetic marks, affecting gene expression patterns associated with normal lymphoid development. This epigenetic dysregulation can drive the expression of genes promoting leukemic transformation and contribute to ALL development. PU.1 plays a crucial role in directing hematopoietic progenitor cells towards myeloid or lymphoid lineages [45]. Dysregulated PU.1 activity may disrupt normal cell fate decisions, leading to the emergence of leukemic progenitor cells with altered differentiation potential. This altered cell fate decision-making can contribute to the development of ALL by promoting the expansion of leukemic clones with self-renewal capabilities and impaired differentiation. Dysregulation of PU.1 can lead to aberrant downstream gene expression, TF activity, signaling pathway activation, epigenetic modifications, and cell fate decisions, ultimately contributing to the development of ALL leukemogenesis [46, 47].

##### Down Syndrome based mutations (GATA1, CRLF2, JAK2, RAS) in ALL Leukemogenesis and in increasing ALL Risk

While PU.1 itself is not located on chromosome 21, the altered genomic context in DS may indirectly affect PU.1 expression through chromatin remodeling or changes in transcriptional regulatory networks [48]. DS-associated alterations in chromatin structure or epigenetic marks may impact the expression of genes involved in regulating PU.1 expression [49]. For example, changes in DNA methylation patterns or histone modifications may affect PU.1 transcriptional activity. GATA1 mutations in DS-ALL have been associated with dysregulated PU.1 expression. GATA1 directly regulates PU.1 expression, and mutations in GATA1 can disrupt its regulatory function, leading to altered PU.1 levels [50]. Dysregulated PU.1 expression due to GATA1 mutations, compounded by potential dosage effects in DS, may disrupt normal hematopoietic differentiation, promoting the development of ALL by altering the balance between myeloid and lymphoid lineage commitment [51]. Dysregulated CRLF2 signaling in DS-ALL can impact PU.1 expression indirectly by altering signaling pathways involved in its regulation. High CRLF2 levels have been associated with decreased PU.1 expression. Altered CRLF2 signaling and reduced PU.1 expression, in the context of DS, may further perturb normal hematopoietic differentiation, contributing to ALL development by promoting aberrant myeloid or lymphoid lineage commitment. JAK2 mutations in DS-ALL activate downstream signaling pathways, including the JAK/STAT pathway, which can modulate PU.1 expression indirectly [52]. Dysregulated JAK2 signaling has been associated with altered PU.1 levels. Aberrant JAK2 signaling and dysregulated PU.1 expression, exacerbated by the genomic context of DS, may disrupt normal hematopoietic differentiation, promoting ALL development by altering the balance between myeloid and lymphoid lineage commitment [53]. Mutations in the RAS pathway, such as activating mutations in RAS genes, can influence PU.1 expression through downstream signaling cascades involved in its regulation. Dysregulated RAS signaling has been associated with altered PU.1 levels [54]. Dysregulated RAS signaling and altered PU.1 expression, in the context of DS, may disrupt normal hematopoietic differentiation, contributing to ALL development by promoting aberrant myeloid or lymphoid lineage commitment. DS-associated mutations in GATA1, CRLF2, JAK2, and RAS may impact PU.1 expression through various mechanisms, including direct regulation, signaling pathway alterations, and epigenetic modifications [55].

#### 3. E2A (TCF3) Transcription Factor

##### Impact of Dysregulations in Disrupting the Cell Fate of Lymphoid Progenitor Cells

E2A, also known as TCF3, is a transcription factor crucial for the developmental biology of Lymphoid Progenitor Cells (CLPs). Dysregulations in E2A expression or function can significantly impact the cell fate of CLPs, potentially leading to the development of Acute Lymphoblastic Leukemia (ALL) [56]. E2A regulates lineage commitment by promoting the differentiation of CLPs into B-cell and T-cell lineages while inhibiting alternative lineage fates. Dysregulated E2A expression or function may lead to aberrant lineage commitment, skewing the balance towards myeloid or other non-lymphoid lineage commitment. This imbalance can result in the expansion of progenitors with altered differentiation potential, predisposing to ALL development [57]. E2A also regulates the expression of genes involved in lymphoid differentiation, ensuring the proper maturation of CLPs into mature B and T lymphocytes. Dysregulated E2A activity may impair lymphoid differentiation, leading to the accumulation of immature or undifferentiated lymphoid progenitors. This block in differentiation can create a pool of precursor cells with self-renewal capabilities, contributing to the leukemic transformation observed in ALL [58]. Additionally, E2A plays a role in regulating cell proliferation, maintaining the balance between proliferation and differentiation in hematopoietic progenitor cells. Dysregulated E2A expression or function may confer a proliferative advantage to CLPs, promoting uncontrolled cell proliferation and the expansion of leukemic clones. This dysregulated proliferation can lead to the accumulation of leukemic blasts characteristic of ALL. Moreover, E2A regulates the expression of genes involved in DNA repair and maintenance of genomic stability. Dysregulated E2A activity may lead to genomic instability, resulting in the accumulation of genetic mutations and chromosomal abnormalities in CLPs [59]. This genomic instability can drive leukemogenesis by promoting the acquisition of additional mutations necessary for ALL development. Furthermore, E2A governs cell fate decisions in hematopoietic progenitors, directing them towards B-cell or T-cell lineages. Dysregulated E2A activity may disrupt normal cell fate decisions, leading to the emergence of leukemic progenitors with altered differentiation potential. This altered cell fate decision-making can contribute to the development of ALL by promoting the expansion of leukemic clones with self-renewal capabilities and impaired differentiation. Dysregulations in E2A expression or function can disrupt normal lymphoid development and predispose Lymphoid Progenitor Cells to ALL development [60].

##### Damage leading to ALL Leukemogenesis

Dysregulation of E2A (TCF3) can lead to the development of Acute Lymphoblastic Leukemia (ALL) by altering downstream genes, transcription factors (TFs), and signaling pathways critical for normal lymphoid development. E2A regulates the expression of genes involved in lymphoid lineage commitment, differentiation, and cell cycle control [61]. Dysregulated E2A activity may result in aberrant expression of downstream genes crucial for lymphoid development, favoring leukemogenesis by promoting proliferation, survival, and self-renewal while suppressing differentiation and apoptosis-related genes. E2A interacts with various TFs to regulate gene expression and cell fate decisions during hematopoiesis. Dysregulated E2A may disrupt interactions with its binding partners, altering TF activity, such as dysregulated activity of TFs involved in lymphoid lineage specification, proliferation, and survival, contributing to dysregulated gene expression patterns observed in ALL. Moreover, E2A integrates signals from various signaling pathways, such as Notch, Wnt, and cytokine signaling, to regulate hematopoietic progenitor cell fate decisions [62]. Dysregulated E2A can perturb signaling pathway activity, promoting abnormal proliferation and differentiation of progenitor cells, contributing to ALL pathogenesis. Furthermore, E2A influences chromatin accessibility and histone modifications, thereby modulating gene expression during hematopoiesis. Dysregulated E2A activity may lead to alterations in chromatin structure and epigenetic marks, driving the expression of genes promoting leukemic transformation and contributing to ALL development [63]. E2A plays a crucial role in directing hematopoietic progenitor cells towards lymphoid lineage commitment and differentiation. Dysregulated E2A activity may disrupt normal cell fate decisions, leading to the emergence of leukemic progenitor cells with altered differentiation potential [64]. This altered cell fate decision-making can contribute to the development of ALL by promoting the expansion of leukemic clones with self-renewal capabilities and impaired differentiation [65].

##### Down Syndrome based mutations (GATA1, CRLF2, JAK2, RAS) in ALL Leukemogenesis and in increasing ALL Risk

E2A (TCF3) is located on chromosome 19, and trisomy 21 in DS results in an additional copy of chromosome 21. While E2A itself is not located on chromosome 21, the altered genomic context in DS may indirectly affect E2A expression through chromatin remodeling or changes in transcriptional regulatory networks [66]. DS-associated alterations in chromatin structure or epigenetic marks may impact the expression of genes involved in regulating E2A (TCF3) expression, such as changes in DNA methylation patterns or histone modifications affecting E2A transcriptional activity. GATA1 mutations in DS-ALL have been associated with dysregulated E2A (TCF3) expression. GATA1 directly regulates E2A expression, and mutations in GATA1 can disrupt its regulatory function, leading to altered E2A levels. Dysregulated E2A (TCF3) expression due to GATA1 mutations, compounded by potential dosage effects in DS, may disrupt normal lymphoid development, promoting ALL development by altering the balance between myeloid and lymphoid lineage commitment [67]. Similarly, dysregulated CRLF2 signaling in DS-ALL can impact E2A (TCF3) expression indirectly by altering signaling pathways involved in its regulation. High CRLF2 levels have been associated with decreased E2A (TCF3) expression, contributing to ALL development by promoting aberrant myeloid or lymphoid lineage commitment. JAK2 mutations in DS-ALL activate downstream signaling pathways, including the JAK/STAT pathway, which can modulate E2A (TCF3) expression indirectly [68]. Dysregulated JAK2 signaling has been associated with altered E2A (TCF3) levels, disrupting normal lymphoid development and promoting ALL development by altering the balance between myeloid and lymphoid lineage commitment. Similarly, mutations in the RAS pathway, such as activating mutations in RAS genes, can influence E2A (TCF3) expression through downstream signaling cascades involved in its regulation [69]. Dysregulated RAS signaling and altered E2A (TCF3) expression in the context of DS may disrupt normal lymphoid development, contributing to ALL development by promoting aberrant myeloid or lymphoid lineage commitment [70]. DS-associated mutations in GATA1, CRLF2, JAK2, and RAS may impact E2A (TCF3) expression through various mechanisms, including direct regulation, signaling pathway alterations, and epigenetic modifications [71, 72].

#### 4. EBF1 Transcription Factor

##### Impact of Dysregulations in Disrupting the Cell Fate of Lymphoid Progenitor Cells

EBF1 (Early B-cell Factor 1) is a transcription factor that plays a crucial role in the developmental biology of Lymphoid Progenitor Cells (CLPs), particularly in B-cell lineage commitment and differentiation. Dysregulations in EBF1 expression or function can significantly impact the cell fate of CLPs, potentially leading to the development of Acute Lymphoblastic Leukemia (ALL) [73]. EBF1 promotes the commitment of CLPs towards the B-cell lineage while suppressing alternative lineage fates, such as myeloid or T-cell lineages. Dysregulated EBF1 expression or function may lead to aberrant lineage commitment, skewing the balance towards alternative lineage fates or impairing normal B-cell development [74]. This imbalance can result in the expansion of progenitors with altered differentiation potential, predisposing to ALL development. EBF1 regulates the expression of genes involved in B-cell differentiation, including those encoding immunoglobulin genes and B-cell-specific surface markers. Dysregulated EBF1 activity may impair B-cell differentiation, leading to the accumulation of immature or undifferentiated B-cell progenitors. This block in differentiation can create a pool of precursor cells with self-renewal capabilities, contributing to the leukemic transformation observed in ALL [75]. EBF1 acts as a regulator of cell proliferation in B-cell progenitors, maintaining the balance between proliferation and differentiation. Dysregulated EBF1 expression or function may confer a proliferative advantage to CLPs, promoting uncontrolled cell proliferation and the expansion of leukemic clones. This dysregulated proliferation can lead to the accumulation of leukemic blasts characteristic of ALL. EBF1 regulates the expression of genes involved in DNA repair and maintenance of genomic stability in developing B-cells [76]. Dysregulated EBF1 activity may lead to genomic instability, resulting in the accumulation of genetic mutations and chromosomal abnormalities in CLPs. This genomic instability can drive leukemogenesis by promoting the acquisition of additional mutations necessary for ALL development. EBF1 governs cell fate decisions in B-cell progenitors, directing them towards B-cell lineage commitment and differentiation [77]. Dysregulated EBF1 activity may disrupt normal cell fate decisions, leading to the emergence of leukemic progenitors with altered differentiation potential. This altered cell fate decision-making can contribute to the development of ALL by promoting the expansion of leukemic clones with self-renewal capabilities and impaired differentiation [78].

##### Damage leading to ALL Leukemogenesis

Dysregulation of EBF1 (Early B-cell Factor 1) can lead to the development of Acute Lymphoblastic Leukemia (ALL) by altering downstream genes, transcription factors (TFs), and signaling pathways critical for normal lymphoid development [79]. EBF1 regulates the expression of genes essential for B-cell lineage commitment, differentiation, and maturation, including those encoding immunoglobulins and B-cell-specific transcription factors. Dysregulated EBF1 activity may result in aberrant expression of downstream genes crucial for B-cell development. This dysregulation can lead to the upregulation of genes promoting proliferation, survival, and self-renewal while suppressing genes involved in differentiation and apoptosis, thereby favoring leukemogenesis [80]. EBF1 interacts with various TFs to regulate gene expression during B-cell development, including Pax5, E2A, and PU.1. Dysregulated EBF1 activity may disrupt interactions with its binding partners, altering TF activity. For example, impaired EBF1 function may lead to dysregulated activity of TFs involved in B-cell lineage commitment, proliferation, and survival, contributing to the dysregulated gene expression patterns observed in ALL [81]. EBF1 integrates signals from various signaling pathways, such as Notch, Wnt, and cytokine signaling, to regulate B-cell fate decisions. Dysregulated EBF1 can perturb signaling pathway activity, leading to aberrant cellular responses. For instance, impaired EBF1 function may dysregulate Notch or Wnt signaling pathways critical for B-cell development, promoting abnormal proliferation and differentiation of progenitor cells and contributing to ALL pathogenesis [82]. EBF1 influences chromatin accessibility and histone modifications, thereby modulating gene expression during B-cell development. Dysregulated EBF1 activity may lead to alterations in chromatin structure and epigenetic marks, affecting gene expression patterns associated with normal B-cell development. This epigenetic dysregulation can drive the expression of genes promoting leukemic transformation and contribute to ALL development. EBF1 plays a crucial role in directing B-cell progenitor cells towards B-cell lineage commitment and differentiation. Dysregulated EBF1 activity may disrupt normal cell fate decisions, leading to the emergence of leukemic progenitor cells with altered differentiation potential [83]. This altered cell fate decision-making can contribute to the development of ALL by promoting the expansion of leukemic clones with self-renewal capabilities and impaired differentiation [84].

##### Down Syndrome based mutations (GATA1, CRLF2, JAK2, RAS) in ALL Leukemogenesis and in increasing ALL Risk

EBF1 is located on chromosomes 5 and 14 in humans, and trisomy 21 in DS results in an additional copy of chromosome 21. While EBF1 itself is not located on chromosome 21, the altered genomic context in DS may indirectly affect EBF1 expression through chromatin remodeling or changes in transcriptional regulatory networks. DS-associated alterations in chromatin structure or epigenetic marks may impact the expression of genes involved in regulating EBF1 expression [85]. For example, changes in DNA methylation patterns or histone modifications may affect EBF1 transcriptional activity. GATA1 mutations in DS-ALL have been associated with dysregulated EBF1 expression. GATA1 directly regulates EBF1 expression, and mutations in GATA1 can disrupt its regulatory function, leading to altered EBF1 levels. Dysregulated CRLF2 signaling in DS-ALL can impact EBF1 expression indirectly by altering signaling pathways involved in its regulation. High CRLF2 levels have been associated with decreased EBF1 expression. Aberrant JAK2 signaling and dysregulated EBF1 expression, exacerbated by the genomic context of DS, may disrupt normal lymphoid development, promoting ALL development by altering the balance between myeloid and lymphoid lineage commitment [86]. Mutations in the RAS pathway, such as activating mutations in RAS genes, can influence EBF1 expression through downstream signaling cascades involved in its regulation. Dysregulated RAS signaling and altered EBF1 expression, in the context of DS, may disrupt normal lymphoid development, contributing to ALL development by promoting aberrant myeloid or lymphoid lineage commitment [87, 88].

#### 5. Notch1 Transcription Factor

##### Impact of Dysregulations in Disrupting the Cell Fate of Lymphoid Progenitor Cells

Notch1 signaling plays a crucial role in the developmental biology of Lymphoid Progenitor Cells (CLPs), particularly in regulating cell fate decisions and lineage commitment [89]. Dysregulations in Notch1 expression or activity can significantly impact the cell fate of CLPs, potentially leading to the development of Acute Lymphoblastic Leukemia (ALL). Notch1 signaling is involved in directing CLPs towards the T-cell lineage by promoting T-cell fate decisions while inhibiting alternative lineage commitments. Dysregulated Notch1 activity may lead to aberrant lineage commitment, skewing the balance towards myeloid or other non-lymphoid lineage commitment [90]. This imbalance can result in the expansion of progenitors with altered differentiation potential, predisposing to ALL development. Notch1 signaling regulates the expression of genes involved in T-cell differentiation, including those encoding T-cell-specific transcription factors and surface markers. Dysregulated Notch1 activity may impair T-cell differentiation, leading to the accumulation of immature or undifferentiated T-cell progenitors [91]. This block in differentiation can create a pool of precursor cells with self-renewal capabilities, contributing to the leukemic transformation observed in ALL. Notch1 signaling also regulates cell proliferation and survival in CLPs, maintaining the balance between proliferation and differentiation. Dysregulated Notch1 expression or activity may confer a proliferative advantage to CLPs, promoting uncontrolled cell proliferation and the expansion of leukemic clones. This dysregulated proliferation can lead to the accumulation of leukemic blasts characteristic of ALL. Furthermore, Notch1 signaling influences genomic stability by regulating cell cycle progression and DNA repair mechanisms in developing T-cells. Dysregulated Notch1 activity may lead to genomic instability, resulting in the accumulation of genetic mutations and chromosomal abnormalities in CLPs [92]. This genomic instability can drive leukemogenesis by promoting the acquisition of additional mutations necessary for ALL development [93].

##### Damage leading to ALL Leukemogenesis

Dysregulation of Notch1 signaling can lead to the development of Acute Lymphoblastic Leukemia (ALL) by altering downstream genes, transcription factors (TFs), and signaling pathways critical for normal lymphoid development. Here’s how damaged Notch1 may switch these downstream elements towards a pathologic state, contributing to ALL leukemogenesis [94]. Notch1 signaling regulates the expression of genes involved in cell fate determination, proliferation, and survival in Lymphoid Progenitor Cells (CLPs). Dysregulated Notch1 activity can lead to aberrant expression of downstream genes, including those promoting cell proliferation, inhibiting apoptosis, and altering differentiation programs [95]. This dysregulated gene expression can contribute to the leukemic transformation of CLPs observed in ALL. Notch1 signaling interacts with various TFs, including HES1, MYC, and GATA3, to regulate gene expression and cell fate decisions. Dysregulated Notch1 activity may disrupt interactions with its TF partners, altering TF activity and gene expression patterns. For example, impaired Notch1 function may lead to dysregulated activity of TFs involved in lymphoid lineage commitment and differentiation, contributing to ALL pathogenesis. Notch1 signaling crosstalks with multiple signaling pathways, such as Wnt, PI3K/AKT, and NF-κB, to regulate various cellular processes. Dysregulated Notch1 signaling can perturb the activity of downstream signaling pathways, leading to aberrant cellular responses [96]. For instance, impaired Notch1 function may dysregulate PI3K/AKT signaling, promoting cell survival and proliferation, which are hallmarks of leukemic cells in ALL. Notch1 signaling influences chromatin modifications and epigenetic regulation of gene expression during lymphoid development. Dysregulated Notch1 activity may lead to alterations in epigenetic marks and chromatin structure, affecting gene expression patterns associated with normal lymphoid development [97]. This epigenetic dysregulation can drive the expression of genes promoting leukemic transformation and contribute to ALL development. Notch1 governs cell fate decisions in CLPs, directing them towards T-cell lineage commitment and differentiation. Dysregulated Notch1 activity may disrupt normal cell fate decisions, leading to the emergence of leukemic progenitors with altered differentiation potential. This altered cell fate decision-making can contribute to the development of ALL by promoting the expansion of leukemic clones with self-renewal capabilities and impaired differentiation [98].

##### Down Syndrome based mutations (GATA1, CRLF2, JAK2, RAS) in ALL Leukemogenesis and in increasing ALL Risk

Notch1 is located on chromosome 9 in humans, and trisomy 21 in Down Syndrome (DS) results in an additional copy of chromosome 21. While Notch1 itself is not located on chromosome 21, the altered genomic context in DS may indirectly affect Notch1 expression through chromatin remodeling or changes in transcriptional regulatory networks [99]. DS-associated alterations in chromatin structure or epigenetic marks may impact the expression of genes involved in regulating Notch1 expression. For example, changes in DNA methylation patterns or histone modifications may affect Notch1 transcriptional activity. GATA1 mutations in DS-ALL have been associated with dysregulated Notch1 expression. GATA1 directly regulates Notch1 expression, and mutations in GATA1 can disrupt its regulatory function, leading to altered Notch1 levels [100]. Dysregulated Notch1 expression due to GATA1 mutations, compounded by potential dosage effects in DS, may disrupt normal lymphoid development, promoting the development of ALL by altering the balance between myeloid and lymphoid lineage commitment. Dysregulated CRLF2 signaling in DS-ALL can impact Notch1 expression indirectly by altering signaling pathways involved in its regulation. High CRLF2 levels have been associated with decreased Notch1 expression [101]. Altered CRLF2 signaling and reduced Notch1 expression, in the context of DS, may further perturb normal lymphoid development, contributing to ALL development by promoting aberrant myeloid or lymphoid lineage commitment. JAK2 mutations in DS-ALL activate downstream signaling pathways, including the JAK/STAT pathway, which can modulate Notch1 expression indirectly. Dysregulated JAK2 signaling has been associated with altered Notch1 levels. Aberrant JAK2 signaling and dysregulated Notch1 expression, exacerbated by the genomic context of DS, may disrupt normal lymphoid development, promoting ALL development by altering the balance between myeloid and lymphoid lineage commitment. Mutations in the RAS pathway, such as activating mutations in RAS genes, can influence Notch1 expression through downstream signaling cascades involved in its regulation. Dysregulated RAS signaling has been associated with altered Notch1 levels [102]. Dysregulated RAS signaling and altered Notch1 expression, in the context of DS, may disrupt normal lymphoid development, contributing to ALL development by promoting aberrant myeloid or lymphoid lineage commitment [103, 104]. DS-associated mutations in GATA1, CRLF2, JAK2, and RAS may impact Notch1 expression through various mechanisms, including direct regulation, signaling pathway alterations, and epigenetic modifications [105, 106].

#### 6. CD34 Surface Markers

##### Impact of Dysregulations in Disrupting the Cell Fate of Lymphoid Progenitor Cells

CD34 is a cell surface glycoprotein expressed on hematopoietic stem cells (HSCs) and early progenitor cells, including Lymphoid Progenitor Cells (CLPs), playing a crucial role in hematopoiesis and stem cell maintenance. Dysregulations in CD34 expression or function can impact CLPs’ cell fate, potentially contributing to Acute Lymphoblastic Leukemia (ALL). CD34’s dysregulation may disrupt CLPs’ cell fate in several ways [107]. CD34 is involved in maintaining stem cell properties like self-renewal and multipotency in HSCs and progenitor cells. Dysregulated CD34 expression or function can alter stem cell properties in CLPs, including aberrant self-renewal or differentiation potential, leading to the accumulation of leukemic progenitor cells with enhanced proliferative capacity and impaired differentiation, thus contributing to ALL development. Additionally, CD34 expression is downregulated as progenitor cells commit to specific hematopoietic lineages, including the lymphoid lineage [108]. Dysregulated CD34 expression or function may disrupt normal lymphoid lineage commitment in CLPs, leading to aberrant differentiation towards lymphoid cells, which can result in the expansion of abnormal lymphoid progenitors predisposed to leukemic transformation, contributing to ALL development. Furthermore, CD34 expression is associated with the regulation of cell survival and proliferation in hematopoietic progenitor cells [109]. Dysregulated CD34 expression or function may confer a survival or proliferative advantage to CLPs, promoting the expansion of leukemic clones, leading to the accumulation of leukemic blasts characteristic of ALL. CD34 also mediates cell-cell and cell-matrix interactions, playing a role in cell adhesion and migration during hematopoiesis. Dysregulated CD34 expression or function may alter the adhesive properties or migratory capacity of CLPs, affecting their localization within the bone marrow microenvironment, which can contribute to the dissemination of leukemic cells and disease progression in ALL. Moreover, CD34 expression has been associated with chemoresistance in leukemic cells. Dysregulated CD34 expression or function may confer resistance to chemotherapy in leukemic progenitor cells, allowing them to evade cytotoxic treatments and persist in the bone marrow niche, which can contribute to disease relapse and poor outcomes in ALL patients [110, 111].

##### Damage leading to ALL Leukemogenesis

Dysregulation of CD34, a cell surface glycoprotein expressed on hematopoietic stem cells and progenitor cells, can influence the development of Acute Lymphoblastic Leukemia (ALL) by impacting downstream genes, transcription factors (TFs), and signaling pathways critical for hematopoietic differentiation and leukemogenesis [112]. CD34 plays a role in regulating the expression of genes involved in hematopoietic differentiation, proliferation, and survival. Dysregulated CD34 expression may lead to aberrant expression of downstream genes that promote leukemogenesis [113]. This dysregulated gene expression can include upregulation of genes involved in cell proliferation (e.g., MYC, Cyclin D1) and inhibition of genes involved in apoptosis or differentiation, contributing to the development and maintenance of leukemic cells in ALL. CD34 may interact with TFs such as GATA2, PU.1, and C/EBPα to regulate gene expression in hematopoietic progenitor cells [114]. Dysregulated CD34 expression may disrupt interactions with TFs involved in normal hematopoiesis, leading to altered TF activity. This dysregulation can result in aberrant expression of downstream target genes that drive leukemic transformation, contributing to ALL pathogenesis. Moreover, CD34 expression influences various signaling pathways, including PI3K/AKT, MAPK/ERK, and JAK/STAT pathways, which regulate hematopoietic cell survival, proliferation, and differentiation [115]. Dysregulated CD34 expression may lead to aberrant activation or inhibition of signaling pathways critical for leukemogenesis. For instance, dysregulated CD34 signaling may activate prosurvival pathways (e.g., PI3K/AKT) while inhibiting pathways promoting apoptosis, thereby promoting the survival and proliferation of leukemic cells in ALL. CD34 expression also influences epigenetic modifications, including DNA methylation and histone acetylation, which regulate gene expression during hematopoietic differentiation [116]. Dysregulated CD34 expression may alter epigenetic modifications in leukemic cells, leading to changes in gene expression patterns that favor leukemogenesis. This dysregulation can promote the silencing of tumor suppressor genes or the activation of oncogenes, contributing to ALL development. Additionally, CD34 expression influences hematopoietic cell fate decisions, including self-renewal, proliferation, and differentiation [117]. Dysregulated CD34 expression may disrupt normal cell fate decisions, leading to the accumulation of leukemic progenitor cells with altered differentiation potential. This altered cell fate decision-making can contribute to the expansion of leukemic clones with self-renewal capabilities and impaired differentiation, driving ALL leukemogenesis [118, 119].

##### Down Syndrome based mutations (GATA1, CRLF2, JAK2, RAS) in ALL Leukemogenesis and in increasing ALL Risk

GATA1 mutations in DS-ALL have been associated with dysregulated CD34 expression. GATA1 directly regulates CD34 expression, and mutations in GATA1 can disrupt its regulatory function, leading to altered CD34 levels [120]. Dysregulated CD34 expression due to GATA1 mutations, compounded by potential dosage effects in DS, may disrupt normal lymphoid development, promoting the development of ALL by altering the balance between myeloid and lymphoid lineage commitment [121]. Moreover, dysregulated CRLF2 signaling in DS-ALL can impact CD34 expression indirectly by altering signaling pathways involved in its regulation. High CRLF2 levels have been associated with altered CD34 expression. Altered CRLF2 signaling and dysregulated CD34 expression, in the context of DS, may perturb normal lymphoid development, contributing to ALL development by promoting aberrant myeloid or lymphoid lineage commitment [122]. Similarly, JAK2 mutations in DS-ALL activate downstream signaling pathways, including the JAK/STAT pathway, which can modulate CD34 expression indirectly. Dysregulated JAK2 signaling has been associated with altered CD34 levels. Aberrant JAK2 signaling and dysregulated CD34 expression, exacerbated by the genomic context of DS, may disrupt normal lymphoid development, promoting ALL development by altering the balance between myeloid and lymphoid lineage commitment. Additionally, mutations in the RAS pathway, such as activating mutations in RAS genes, can influence CD34 expression through downstream signaling cascades involved in its regulation [123]. Dysregulated RAS signaling has been associated with altered CD34 levels. Dysregulated RAS signaling and altered CD34 expression, in the context of DS, may disrupt normal lymphoid development, contributing to ALL development by promoting aberrant myeloid or lymphoid lineage commitment [124]. DS-associated mutations in GATA1, CRLF2, JAK2, and RAS may impact CD34 expression through various mechanisms, including direct regulation, signaling pathway alterations, and epigenetic modifications [125, 126].

#### 7. CD38 Surface Marker

##### Impact of Dysregulations in Disrupting the Cell Fate of Lymphoid Progenitor Cells

CD38, a transmembrane glycoprotein, is expressed on various hematopoietic cells, including Lymphoid Progenitor Cells (CLPs), where it plays essential roles in cell signaling, adhesion, and differentiation. Dysregulation of CD38 expression or function can impact the cell fate of CLPs and potentially contribute to the development of Acute Lymphoblastic Leukemia (ALL) [127]. CD38 acts as a multifunctional enzyme involved in calcium signaling and the generation of second messengers, such as cyclic ADP-ribose (cADPR) and nicotinic acid adenine dinucleotide phosphate (NAADP). Dysregulated CD38 expression or function may lead to altered intracellular signaling pathways in CLPs, affecting critical processes such as cell proliferation, differentiation, and survival [128]. Aberrant signaling may disrupt normal lymphoid development and contribute to the expansion of leukemic progenitor cells characteristic of ALL. Moreover, CD38 participates in cell adhesion and migration processes by interacting with extracellular matrix components and cell surface receptors. Dysregulated CD38 expression or function may disrupt cell adhesion and migration properties of CLPs, affecting their localization within the bone marrow microenvironment [129]. Altered adhesion and migration can impact interactions with stromal cells and niche factors critical for normal hematopoiesis, potentially promoting the dissemination and survival of leukemic cells in ALL. CD38 expression is associated with specific immunophenotypic markers used for identifying hematopoietic cell subsets, including CLPs. Dysregulated CD38 expression may alter the immunophenotypic profile of CLPs, making them more or less susceptible to leukemic transformation [130]. Changes in CD38 expression levels or localization may affect the differentiation potential or functional properties of CLPs, contributing to the development of ALL. Furthermore, CD38 expression influences interactions between hematopoietic cells and the bone marrow microenvironment, including stromal cells, endothelial cells, and immune cells. Dysregulated CD38 expression or function may disrupt normal interactions between CLPs and the hematopoietic microenvironment. Alterations in cell-cell communication or signaling pathways within the niche may create a permissive environment for leukemic transformation and expansion, facilitating ALL development [131]. CD38 expression is associated with modulation of apoptosis and cell survival pathways in hematopoietic cells. Dysregulated CD38 expression or function may affect apoptotic signaling pathways in CLPs, leading to dysregulated cell survival and increased resistance to apoptosis. This dysregulation may contribute to the accumulation of leukemic progenitor cells and the progression of ALL [132].

##### Damage leading to ALL Leukemogenesis

CD38 regulates the expression of genes involved in cell proliferation, differentiation, and survival through its enzymatic activity and signaling functions. Dysregulated CD38 expression or function can lead to aberrant expression of downstream genes associated with leukemogenesis [133]. This dysregulation may include upregulation of oncogenes (e.g., MYC, BCL2) or downregulation of tumor suppressor genes, promoting uncontrolled proliferation, impaired differentiation, and increased survival of leukemic cells in ALL. Moreover, CD38 can modulate the activity of TFs involved in hematopoietic development and leukemogenesis, such as NF-κB, STATs, and AP-1. Dysregulated CD38 expression may disrupt the activity of TFs critical for normal hematopoiesis, leading to altered gene expression profiles favoring leukemic transformation [134]. This dysregulation can promote the expression of TFs associated with cell proliferation, survival, and resistance to apoptosis, contributing to ALL pathogenesis. Furthermore, CD38 influences various signaling pathways, including calcium signaling, MAPK/ERK, PI3K/AKT, and NF-κB pathways, which regulate cell fate decisions in hematopoietic cells. Dysregulated CD38 expression can perturb signaling pathways involved in cell proliferation, survival, and differentiation, promoting the development of leukemic phenotypes. For example, dysregulated CD38 signaling may activate prosurvival pathways while inhibiting pathways promoting apoptosis, driving the expansion and survival of leukemic cells in ALL [135]. Additionally, CD38 activity influences epigenetic modifications, including histone acetylation and DNA methylation, which regulate gene expression patterns during hematopoietic differentiation. Dysregulated CD38 expression or function may alter epigenetic modifications associated with leukemogenesis, leading to changes in gene expression profiles favoring leukemic transformation. This dysregulation can promote the silencing of tumor suppressor genes or the activation of oncogenes, contributing to the initiation and progression of ALL [136]. Moreover, CD38 plays a role in regulating cell fate decisions, including proliferation, differentiation, and apoptosis, in hematopoietic progenitor cells. Dysregulated CD38 expression may disrupt normal cell fate decisions, leading to the expansion of leukemic progenitor cells with altered differentiation potential and increased self-renewal capacity. This dysregulation can contribute to the development and maintenance of leukemic phenotypes observed in ALL [137].

##### Down Syndrome based mutations (GATA1, CRLF2, JAK2, RAS) in ALL Leukemogenesis and in increasing ALL Risk

While CD38 is not located on chromosome 21, the altered genomic context in Down Syndrome (DS) may indirectly affect CD38 expression through chromatin remodeling or changes in transcriptional regulatory networks. DS-associated alterations in chromatin structure or epigenetic marks may impact the expression of genes involved in regulating CD38 expression [138]. For example, changes in DNA methylation patterns or histone modifications may affect CD38 transcriptional activity. GATA1 mutations in DS-ALL have been associated with dysregulated CD38 expression. GATA1 directly regulates CD38 expression, and mutations in GATA1 can disrupt its regulatory function, leading to altered CD38 levels. Dysregulated CD38 expression due to GATA1 mutations, compounded by potential dosage effects in DS, may disrupt normal lymphoid development, promoting the development of ALL by altering the balance between myeloid and lymphoid lineage commitment [139]. Moreover, dysregulated CRLF2 signaling in DS-ALL can impact CD38 expression indirectly by altering signaling pathways involved in its regulation. High CRLF2 levels have been associated with altered CD38 expression. Altered CRLF2 signaling and dysregulated CD38 expression, in the context of DS, may perturb normal lymphoid development, contributing to ALL development by promoting aberrant myeloid or lymphoid lineage commitment [140]. Additionally, JAK2 mutations in DS-ALL activate downstream signaling pathways, including the JAK/STAT pathway, which can modulate CD38 expression indirectly. Dysregulated JAK2 signaling has been associated with altered CD38 levels. Aberrant JAK2 signaling and dysregulated CD38 expression, exacerbated by the genomic context of DS, may disrupt normal lymphoid development, promoting ALL development by altering the balance between myeloid and lymphoid lineage commitment. Furthermore, mutations in the RAS pathway, such as activating mutations in RAS genes, can influence CD38 expression through downstream signaling cascades involved in its regulation [141, 142]. Dysregulated RAS signaling has been associated with altered CD38 levels. Dysregulated RAS signaling and altered CD38 expression, in the context of DS, may disrupt normal lymphoid development, contributing to ALL development by promoting aberrant myeloid or lymphoid lineage commitment [143].

#### 8. CD10 Surface Marker

##### Impact of Dysregulations in Disrupting the Cell Fate of Lymphoid Progenitor Cells

CD10, also known as common acute lymphoblastic leukemia antigen (CALLA), is a cell surface metalloproteinase expressed on early lymphoid progenitor cells, including Lymphoid Progenitor Cells (CLPs). It plays a crucial role in lymphoid development and differentiation [144]. Dysregulation of CD10 expression or function can impact the cell fate of CLPs and potentially contribute to the development of Acute Lymphoblastic Leukemia (ALL). Dysregulated CD10 expression may lead to aberrant differentiation of CLPs, resulting in the accumulation of immature lymphoid cells or lymphoid progenitors with blocked or arrested differentiation pathways. This disruption in normal differentiation processes can contribute to the development of ALL by promoting the expansion of leukemic progenitor cells [145]. Moreover, CD10 functions as a signaling molecule involved in cell adhesion, migration, and proliferation through its proteolytic activity. Dysregulated CD10 expression or function may perturb signaling pathways critical for lymphoid development and homeostasis. Aberrant CD10 signaling may lead to enhanced cell proliferation, survival, or migration, contributing to the expansion and dissemination of leukemic cells in ALL. Additionally, CD10 expression has been associated with modulation of apoptosis and cell survival pathways in hematopoietic cells. Dysregulated CD10 expression may affect apoptotic signaling pathways in CLPs, leading to dysregulated cell survival and increased resistance to apoptosis [146]. This dysregulation may contribute to the accumulation of leukemic progenitor cells and the progression of ALL. Furthermore, CD10 is involved in chemotaxis and cell migration processes by cleaving chemokines and extracellular matrix components. Dysregulated CD10 expression may alter the chemotactic response of CLPs to specific microenvironmental cues within the bone marrow niche. This altered migratory behavior can affect the homing and retention of leukemic cells within the bone marrow microenvironment, facilitating disease progression in ALL. Additionally, CD10 expression is associated with regulatory functions in the immune system, including modulation of antigen presentation and T-cell activation [147]. Dysregulated CD10 expression may disrupt immune regulatory mechanisms within the bone marrow microenvironment, leading to dysregulated immune responses or immune evasion strategies by leukemic cells. This dysregulation can contribute to disease progression and immune escape in ALL [148, 149].

##### Damage leading to ALL Leukemogenesis

CD10 regulates the expression of genes involved in cell differentiation, proliferation, and survival through its proteolytic activity and signaling functions. Dysregulated CD10 expression can lead to aberrant expression of downstream genes associated with leukemogenesis [150]. This dysregulation may include upregulation of oncogenes (e.g., MYC, BCL2) or downregulation of tumor suppressor genes, promoting uncontrolled proliferation, impaired differentiation, and increased survival of leukemic cells in ALL [151]. Moreover, CD10 activity influences the activity of TFs involved in hematopoietic development and leukemogenesis, such as NF-κB, STATs, and AP-1. Dysregulated CD10 expression may disrupt the activity of TFs critical for normal hematopoiesis, leading to altered gene expression profiles favoring leukemic transformation [152]. This dysregulation can promote the expression of TFs associated with cell proliferation, survival, and resistance to apoptosis, contributing to ALL pathogenesis. Additionally, CD10 participates in cell signaling pathways involved in cell adhesion, migration, and proliferation through its proteolytic activity and interactions with other cell surface molecules. Dysregulated CD10 expression can perturb signaling pathways critical for cell fate decisions in hematopoietic cells. Aberrant CD10 signaling may lead to enhanced cell proliferation, survival, or migration, contributing to the expansion and dissemination of leukemic cells in ALL. Furthermore, CD10 activity influences epigenetic modifications, including histone cleavage and DNA methylation, which regulate gene expression patterns during hematopoietic differentiation. Dysregulated CD10 expression or function may alter epigenetic modifications associated with leukemogenesis, leading to changes in gene expression profiles favoring leukemic transformation [153]. This dysregulation can promote the silencing of tumor suppressor genes or the activation of oncogenes, contributing to the initiation and progression of ALL. Moreover, CD10 plays a role in regulating cell fate decisions, including proliferation, differentiation, and apoptosis, in hematopoietic progenitor cells [154]. Dysregulated CD10 expression may disrupt normal cell fate decisions, leading to the expansion of leukemic progenitor cells with altered differentiation potential and increased self-renewal capacity. This dysregulation can contribute to the development and maintenance of leukemic phenotypes observed in ALL [155].

##### Down Syndrome based mutations (GATA1, CRLF2, JAK2, RAS) in ALL Leukemogenesis and in increasing ALL Risk

Alterations in the genomic context in Down Syndrome (DS) may indirectly affect CD10 expression through chromatin remodeling or changes in transcriptional regulatory networks, despite CD10 not being located on chromosome 21 [156]. DS-associated alterations in chromatin structure or epigenetic marks may impact the expression of genes involved in regulating CD10 expression. For example, changes in DNA methylation patterns or histone modifications may affect CD10 transcriptional activity [157]. Moreover, GATA1 mutations in DS-ALL have been associated with dysregulated CD10 expression. GATA1 directly regulates CD10 expression, and mutations in GATA1 can disrupt its regulatory function, leading to altered CD10 levels. Dysregulated CD10 expression due to GATA1 mutations, compounded by potential dosage effects in DS, may disrupt normal lymphoid development, promoting the development of ALL by altering the balance between myeloid and lymphoid lineage commitment. Additionally, dysregulated CRLF2 signaling in DS-ALL can impact CD10 expression indirectly by altering signaling pathways involved in its regulation. Elevated CRLF2 levels have been associated with altered CD10 expression. Altered CRLF2 signaling and dysregulated CD10 expression, in the context of DS, may perturb normal lymphoid development, contributing to ALL development by promoting aberrant myeloid or lymphoid lineage commitment. Furthermore, JAK2 mutations in DS-ALL activate downstream signaling pathways, including the JAK/STAT pathway, which can modulate CD10 expression indirectly [158]. Dysregulated JAK2 signaling has been associated with altered CD10 levels. Aberrant JAK2 signaling and dysregulated CD10 expression, exacerbated by the genomic context of DS, may disrupt normal lymphoid development, promoting ALL development by altering the balance between myeloid and lymphoid lineage commitment. Moreover, mutations in the RAS pathway, such as activating mutations in RAS genes, can influence CD10 expression through downstream signaling cascades involved in its regulation. Dysregulated RAS signaling has been associated with altered CD10 levels. Dysregulated RAS signaling and altered CD10 expression, in the context of DS, may disrupt normal lymphoid development, contributing to ALL development by promoting aberrant myeloid or lymphoid lineage commitment [159].

#### 9. CD7 Surface Marker

##### Impact of Dysregulations in Disrupting the Cell Fate of Lymphoid Progenitor Cells

CD7, typically expressed on immature T-cell progenitors, plays a crucial role in early T-cell development, differentiation, and maturation [160]. Dysregulated CD7 expression may lead to aberrant differentiation of Lymphoid Progenitor Cells (CLPs), resulting in the accumulation of immature lymphoid cells or lymphoid progenitors with blocked or arrested differentiation pathways. This disruption in normal differentiation processes can contribute to the development of Acute Lymphoblastic Leukemia (ALL) by promoting the expansion of leukemic progenitor cells [161]. Moreover, CD7 functions as a signaling molecule involved in cell adhesion, activation, and survival of T cells, interacting with various signaling pathways, including the PI3K/AKT and MAPK pathways. Dysregulated CD7 expression or function may perturb signaling pathways critical for lymphoid development and homeostasis. Aberrant CD7 signaling may lead to enhanced cell proliferation, survival, or migration, contributing to the expansion and dissemination of leukemic cells in ALL [162]. Furthermore, CD7 expression has been associated with the modulation of apoptosis and cell survival pathways in T cells. Dysregulated CD7 expression may affect apoptotic signaling pathways in CLPs, leading to dysregulated cell survival and increased resistance to apoptosis. This dysregulation may contribute to the accumulation of leukemic progenitor cells and the progression of ALL [163]. Additionally, CD7 is involved in T-cell homing and migration processes by interacting with chemokines and adhesion molecules. Dysregulated CD7 expression may alter the chemotactic response of CLPs to specific microenvironmental cues within the bone marrow niche. This altered migratory behavior can affect the homing and retention of leukemic cells within the bone marrow microenvironment, facilitating disease progression in ALL. Furthermore, CD7 expression is associated with regulatory functions in the immune system, including modulation of T-cell activation and cytokine production [164]. Dysregulated CD7 expression may disrupt immune regulatory mechanisms within the bone marrow microenvironment, leading to dysregulated immune responses or immune evasion strategies by leukemic cells. This dysregulation can contribute to disease progression and immune escape in ALL [165].

##### Damage leading to ALL Leukemogenesis

CD7 regulates the expression of genes involved in cell differentiation, proliferation, and survival through its interactions with intracellular signaling pathways and transcriptional regulators. Dysregulated CD7 expression can lead to aberrant expression of downstream genes associated with leukemogenesis [166]. This dysregulation may include upregulation of oncogenes (e.g., MYC, BCL2) or downregulation of tumor suppressor genes, promoting uncontrolled proliferation, impaired differentiation, and increased survival of leukemic cells in Acute Lymphoblastic Leukemia (ALL) [167]. Moreover, CD7 activity influences the activity of transcription factors (TFs) involved in hematopoietic development and leukemogenesis, such as NF-κB, STATs, and AP-1. Dysregulated CD7 expression may disrupt the activity of TFs critical for normal hematopoiesis, leading to altered gene expression profiles favoring leukemic transformation [168]. This dysregulation can promote the expression of TFs associated with cell proliferation, survival, and resistance to apoptosis, contributing to ALL pathogenesis. Additionally, CD7 participates in cell signaling pathways involved in cell adhesion, migration, and proliferation through its interactions with various intracellular signaling molecules, including kinases and phosphatases [169]. Dysregulated CD7 expression can perturb signaling pathways critical for cell fate decisions in hematopoietic cells. Aberrant CD7 signaling may lead to enhanced cell proliferation, survival, or migration, contributing to the expansion and dissemination of leukemic cells in ALL. Furthermore, CD7 activity influences epigenetic modifications, including histone acetylation and DNA methylation, which regulate gene expression patterns during hematopoietic differentiation. Dysregulated CD7 expression or function may alter epigenetic modifications associated with leukemogenesis, leading to changes in gene expression profiles favoring leukemic transformation [170]. This dysregulation can promote the silencing of tumor suppressor genes or the activation of oncogenes, contributing to the initiation and progression of ALL. Finally, CD7 plays a role in regulating cell fate decisions, including proliferation, differentiation, and apoptosis, in hematopoietic progenitor cells [171]. Dysregulated CD7 expression may disrupt normal cell fate decisions, leading to the expansion of leukemic progenitor cells with altered differentiation potential and increased self-renewal capacity. This dysregulation can contribute to the development and maintenance of leukemic phenotypes observed in ALL [172].

##### Down Syndrome based mutations (GATA1, CRLF2, JAK2, RAS) in ALL Leukemogenesis and in increasing ALL Risk

While CD7 is not located on chromosome 21, alterations in the genomic context in Down Syndrome (DS) may indirectly affect CD7 expression through chromatin remodeling or changes in transcriptional regulatory networks. DS-associated alterations in chromatin structure or epigenetic marks may impact the expression of genes involved in regulating CD7 expression [173, 174]. Changes in DNA methylation patterns or histone modifications may affect CD7 transcriptional activity. GATA1 mutations in DS-ALL have been associated with dysregulated CD7 expression. GATA1 directly regulates CD7 expression, and mutations in GATA1 can disrupt its regulatory function, leading to altered CD7 levels [175]. Dysregulated CD7 expression due to GATA1 mutations, compounded by potential dosage effects in DS, may disrupt normal lymphoid development, promoting the development of ALL by altering the balance between myeloid and lymphoid lineage commitment. Dysregulated CRLF2 signaling in DS-ALL can impact CD7 expression indirectly by altering signaling pathways involved in its regulation [176] Elevated CRLF2 levels have been associated with altered CD7 expression. Altered CRLF2 signaling and dysregulated CD7 expression, in the context of DS, may perturb normal lymphoid development, contributing to ALL development by promoting aberrant myeloid or lymphoid lineage commitment [177]. JAK2 mutations in DS-ALL activate downstream signaling pathways, including the JAK/STAT pathway, which can modulate CD7 expression indirectly. Dysregulated JAK2 signaling has been associated with altered CD7 levels [178]. Aberrant JAK2 signaling and dysregulated CD7 expression, exacerbated by the genomic context of DS, may disrupt normal lymphoid development, promoting ALL development by altering the balance between myeloid and lymphoid lineage commitment [179]. Mutations in the RAS pathway, such as activating mutations in RAS genes, can influence CD7 expression through downstream signaling cascades involved in its regulation. Dysregulated RAS signaling has been associated with altered CD7 levels. Dysregulated RAS signaling and altered CD7 expression, in the context of DS, may disrupt normal lymphoid development, contributing to ALL development by promoting aberrant myeloid or lymphoid lineage commitment [180].

#### 10. CD45RA Surface Marker

##### Impact of Dysregulations in Disrupting the Cell Fate of Lymphoid Progenitor Cells

CD45RA expression marks early stages of lymphoid progenitor differentiation, gradually decreasing as cells mature. Dysregulated CD45RA expression disrupts CLP differentiation, potentially causing lineage commitment abnormalities or differentiation blockades. This disruption fosters the accumulation of immature or undifferentiated cells, increasing susceptibility to leukemic transformation [181]. CD45RA is crucial for regulating signaling pathways essential in T and B lymphocyte development. Dysregulated CD45RA expression impairs the development of functional immune cells from CLPs, leading to deficiencies in mature lymphoid cell populations. This immune impairment creates an environment conducive to the proliferation and survival of leukemic cells in ALL. Through its phosphatase activity, CD45RA modulates signaling pathways governing cell proliferation, survival, and differentiation [182]. Dysregulated CD45RA expression disrupts these pathways critical for lymphoid progenitor cell fate decisions, potentially promoting aberrant cell proliferation, survival, or differentiation conducive to leukemic transformation and ALL development. CD45RA’s influence on apoptotic signaling pathways regulates cell survival and death decisions [183]. Dysregulated CD45RA expression may alter apoptotic signaling in CLPs, increasing resistance to apoptosis and contributing to the accumulation and progression of leukemic progenitor cells in ALL. CD45RA’s role in mediating interactions between lymphoid progenitor cells and their microenvironment is essential [184]. Dysregulated CD45RA expression disrupts these interactions, leading to changes in cell adhesion, migration, and survival signals. This disruption promotes the survival and expansion of leukemic cells within the bone marrow niche, facilitating ALL progression [185].

##### Damage leading to ALL Leukemogenesis

CD45RA, a key regulator of cell differentiation, proliferation, and survival, modulates downstream gene expression by influencing signaling pathways and transcriptional regulators [186, 187]. Dysregulation of CD45RA expression can lead to aberrant expression of genes associated with leukemogenesis, promoting uncontrolled proliferation, impaired differentiation, and increased survival of leukemic cells in ALL [188]. In normal hematopoietic development, CD45RA activity influences the function of transcription factors (TFs) such as NF-κB, STATs, and AP-1. Dysregulated CD45RA expression may disrupt TF activity critical for hematopoiesis, favoring leukemic transformation by promoting the expression of TFs associated with cell proliferation, survival, and resistance to apoptosis. Participating in cell signaling pathways involved in adhesion, migration, and proliferation, CD45RA interacts with various intracellular signaling molecules [189]. Dysregulated CD45RA expression can perturb these pathways critical for cell fate decisions, potentially enhancing leukemic cell proliferation, survival, or migration in ALL. CD45RA’s involvement in epigenetic regulation, including histone acetylation and DNA methylation, influences gene expression patterns during hematopoietic differentiation [190]. Dysregulated CD45RA expression or function may alter epigenetic modifications associated with leukemogenesis, contributing to the activation of oncogenes or the silencing of tumor suppressor genes in ALL initiation and progression. Regarding cell fate decisions, CD45RA regulates proliferation, differentiation, and apoptosis in hematopoietic progenitor cells. Dysregulated CD45RA expression disrupts these decisions, leading to the expansion of leukemic progenitor cells with altered differentiation potential and increased self-renewal capacity, thereby contributing to the development and maintenance of leukemic phenotypes observed in ALL [191, 192].

##### Down Syndrome based mutations (GATA1, CRLF2, JAK2, RAS) in ALL Leukemogenesis and in increasing ALL Risk

Down Syndrome (DS) can indirectly affect CD45RA expression, despite its location not being on chromosome 21. This impact may occur through chromatin remodeling or changes in transcriptional regulatory networks [193]. Additionally, DS-associated alterations in chromatin structure or epigenetic marks may influence CD45RA expression by affecting the activity of genes involved in its regulation. Modifications in DNA methylation patterns or histone modifications can potentially impact CD45RA transcriptional activity. GATA1 mutations found in DS-ALL are linked to dysregulated CD45RA expression. GATA1, directly or indirectly regulating CD45RA expression, can have its function disrupted by mutations, leading to altered CD45RA levels [194]. This dysregulation, combined with potential dosage effects in DS, may interfere with normal lymphoid development, ultimately promoting ALL by shifting the balance between myeloid and lymphoid lineage commitment. Dysregulated CRLF2 signaling in DS-ALL can indirectly influence CD45RA expression by altering signaling pathways involved in its regulation. Elevated levels of CRLF2 have been associated with changes in CD45RA expression. This altered signaling, along with dysregulated CD45RA expression, in the context of DS, can disrupt normal lymphoid development, potentially contributing to ALL by promoting aberrant myeloid or lymphoid lineage commitment [195]. JAK2 mutations in DS-ALL activate downstream signaling pathways, including the JAK/STAT pathway, which can indirectly modulate CD45RA expression. Dysregulated JAK2 signaling has been correlated with altered CD45RA levels. This aberrant signaling, coupled with dysregulated CD45RA expression, may disrupt normal lymphoid development, thereby promoting ALL by altering the balance between myeloid and lymphoid lineage commitment [196]. Mutations in the RAS pathway, such as activating mutations in RAS genes, can influence CD45RA expression through downstream signaling cascades. Dysregulated RAS signaling has been associated with altered CD45RA levels. In the context of DS, dysregulated RAS signaling and altered CD45RA expression may disrupt normal lymphoid development, potentially contributing to ALL by promoting aberrant myeloid or lymphoid lineage commitment [197].

#### 11. FLT3 (CD135) Surface Marker

##### Impact of Dysregulations in Disrupting the Cell Fate of Lymphoid Progenitor Cells

FLT3 signaling plays a crucial role in regulating the proliferation of lymphoid progenitor cells (LPCs) in response to cytokines such as FLT3 ligand (FL). When FLT3 is aberrantly activated or overexpressed due to mutations or gene amplification, it can lead to excessive proliferation of LPCs, contributing to the expansion of leukemic precursor cells in ALL [198]. Additionally, FLT3 signaling influences the differentiation of LPCs into various hematopoietic lineages, including lymphoid cells. Dysregulated FLT3 signaling may disrupt normal LPC differentiation, resulting in the accumulation of immature or undifferentiated cells with a high proliferative capacity, ultimately blocking the maturation of lymphoid progenitors and contributing to the development of ALL. FLT3 signaling also plays a role in promoting cell survival by inhibiting apoptosis in LPCs [199]. Dysregulated FLT3 activity, particularly as a result of activating mutations, can confer anti-apoptotic properties to LPCs, enhancing their survival and resistance to cell death signals. This dysregulation may contribute to leukemogenesis by enabling the survival and expansion of leukemic cells in ALL [200]. Moreover, FLT3 signaling regulates the migration and homing of LPCs within the bone marrow and other hematopoietic tissues. Dysregulated FLT3 expression or activity may enhance the migratory capacity of LPCs, facilitating their dissemination to extramedullary sites and potentially contributing to leukemic infiltration of organs outside the bone marrow, a characteristic feature of ALL. Furthermore, FLT3 signaling mediates interactions between LPCs and the bone marrow microenvironment, including stromal cells and extracellular matrix components [201]. Dysregulated FLT3 activity may alter these interactions, leading to changes in cell adhesion, cytokine responsiveness, and niche interactions. This dysregulation can create a permissive microenvironment for leukemic precursor cells, promoting their survival and proliferation in ALL [202].

##### Damage leading to ALL Leukemogenesis

FLT3 activation is essential in promoting cell proliferation through downstream signaling pathways like RAS/MAPK and PI3K/AKT pathways [203]. Mutations or overexpression of FLT3 can result in the constitutive activation of these proliferative pathways, fostering uncontrolled cell proliferation characteristic of ALL. Furthermore, FLT3 activation plays a critical role in cell survival by inhibiting apoptotic pathways, such as through the activation of anti-apoptotic proteins like Bcl-2 [204]. Dysregulated FLT3 signaling can disrupt the expression or activity of apoptotic regulators, impairing apoptosis, and enhancing cell survival, thus contributing to leukemogenesis in ALL. In normal physiology, FLT3 signaling regulates hematopoietic stem and progenitor cell differentiation into mature blood cell lineages. However, dysregulated FLT3 activity can disturb normal differentiation pathways, leading to the accumulation of immature progenitor cells and impaired differentiation into mature lymphoid cells, a characteristic hallmark of ALL [205]. FLT3 signaling is involved in regulating hematopoietic stem cell self-renewal and maintenance. Dysregulated FLT3 signaling may activate stemness pathways, resulting in the expansion of leukemic stem cells with enhanced self-renewal capacity, thus contributing to the maintenance and progression of ALL. Moreover, FLT3 activation can influence the expression of genes involved in cell fate decisions, proliferation, and survival through the modulation of transcription factor (TF) activity [206]. Dysregulated FLT3 signaling may lead to altered TF activity, culminating in the aberrant expression of genes associated with leukemogenesis, including oncogenes and tumor suppressors, thereby driving the pathogenesis of ALL. Additionally, FLT3 signaling mediates interactions between leukemic cells and the bone marrow microenvironment, influencing cell adhesion, migration, and survival. Dysregulated FLT3 signaling may perturb these interactions, resulting in changes in the bone marrow niche that support leukemic cell survival, proliferation, and chemoresistance, thus contributing to ALL progression [207, 208].

##### Down Syndrome based mutations (GATA1, CRLF2, JAK2, RAS) in ALL Leukemogenesis and in increasing ALL Risk

FLT3, though not situated on chromosome 21, may experience indirect effects on its expression in Down Syndrome (DS) due to alterations in the genomic context. These alterations could manifest through chromatin remodeling or modifications in transcriptional regulatory networks [209]. Epigenetic changes linked with DS, like modifications in chromatin structure or epigenetic marks, might influence the expression of genes involved in regulating FLT3. Shifts in DNA methylation patterns or histone modifications could consequently impact FLT3 transcriptional activity [210]. GATA1 mutations observed in DS-associated acute lymphoblastic leukemia (ALL) have been linked with disrupted FLT3 expression. GATA1, whether directly or indirectly, regulates FLT3 expression, and mutations in GATA1 could interfere with its regulatory function, resulting in altered FLT3 levels. Such dysregulation, especially in the context of DS, might disturb normal lymphoid development, promoting ALL by altering FLT3-mediated signaling pathways crucial for proliferation, survival, and differentiation [211]. Furthermore, dysregulated CRLF2 signaling in DS-ALL could indirectly influence FLT3 expression by modifying signaling pathways involved in FLT3 regulation. Elevated CRLF2 levels have been correlated with altered FLT3 expression, contributing to aberrant FLT3-mediated signaling and leukemic transformation within the framework of DS, thus disrupting normal lymphoid development and contributing to ALL [212]. Likewise, JAK2 mutations in DS-ALL activate downstream signaling pathways, such as the JAK/STAT pathway, indirectly modulating FLT3 expression. Dysregulated JAK2 signaling has been associated with altered FLT3 levels, exacerbating abnormal FLT3-mediated signaling and leukemic transformation. These aberrations, intensified by the genomic context of DS, may disrupt normal lymphoid development, further contributing to ALL [213]. Moreover, mutations in the RAS pathway, like activating mutations in RAS genes, could influence FLT3 expression through downstream signaling cascades involved in its regulation [214]. Dysregulated RAS signaling has been linked with altered FLT3 levels, disrupting normal lymphoid development and contributing to ALL by promoting aberrant FLT3-mediated signaling and leukemic transformation, particularly in the context of DS [215].

#### 12. Interleukin-7

##### Impact of Dysregulations in Disrupting the Cell Fate of Lymphoid Progenitor Cells

IL-7, a cytokine critical for the development and survival of Lymphoid Progenitor Cells (LPCs) and the regulation of lymphoid cell fate decisions, can become dysregulated, potentially contributing to Acute Lymphoblastic Leukemia (ALL) [216]. Dysregulation of IL-7 signaling may disrupt normal lymphoid development, paving the way for several pathways through which LPC fate can be altered towards ALL. In normal conditions, IL-7 signaling supports LPC survival and proliferation by activating pathways like PI3K/AKT and JAK/STAT. However, aberrant IL-7 signaling, possibly due to overproduction or dysregulated receptor expression, can result in excessive LPC survival and proliferation [217]. This creates a pool of pre-leukemic cells with heightened proliferation potential, a characteristic feature of ALL. Moreover, IL-7 is crucial for promoting LPC differentiation into various lymphoid lineages. Dysregulated IL-7 signaling could disrupt this process, leading to the accumulation of undifferentiated or partially differentiated cells with blocked differentiation pathways. This accumulation of immature lymphoid blasts could contribute to the development of ALL [218]. Additionally, IL-7 signaling influences LPC self-renewal and maintenance of the lymphoid progenitor pool. Dysregulation in this pathway might enhance LPC self-renewal capacity, fostering the expansion of leukemic stem cells with increased potential for long-term survival and propagation of leukemic clones in ALL. Furthermore, IL-7 promotes LPC survival by inhibiting apoptosis through the regulation of Bcl-2 family proteins [219]. Dysregulated IL-7 signaling could disturb apoptotic pathways, increasing the survival of pre-leukemic LPCs and rendering them resistant to cell death signals, thereby contributing to leukemic transformation and maintenance in ALL. Lastly, IL-7 influences LPC interactions with the bone marrow microenvironment, including stromal cells and extracellular matrix components. Dysregulated IL-7 signaling might alter these interactions, resulting in changes in LPC adhesion, migration, and responsiveness to microenvironmental cues. Such alterations can create a permissive niche for the expansion and survival of leukemic cells in ALL [220].

##### Damage leading to ALL Leukemogenesis

IL-7 signaling plays a crucial role in promoting cell proliferation and survival in lymphoid progenitor cells through activation of the JAK/STAT pathway. In normal conditions, IL-7 activates this pathway, facilitating controlled cell proliferation and survival [221]. However, dysregulated IL-7 signaling, possibly due to overproduction of IL-7 or aberrant receptor expression, can result in constitutive activation of the JAK/STAT pathway, fostering uncontrolled cell proliferation, a characteristic feature of ALL. Moreover, IL-7 promotes the differentiation of lymphoid progenitor cells into mature lymphocytes by activating various transcription factors (TFs), including STAT5. Dysregulated IL-7 signaling may inhibit the expression or activity of these TFs critical for lymphoid differentiation [222]. This inhibition can lead to the accumulation of undifferentiated or partially differentiated cells, which is often observed in ALL. Additionally, IL-7 signaling activates PI3K/AKT and Bcl-2 family proteins, promoting cell survival and inhibiting apoptosis. Dysregulated IL-7 signaling can enhance cell survival by upregulating anti-apoptotic proteins and suppressing pro-apoptotic factors. This enhancement in cell survival contributes to the survival and expansion of leukemic cells in ALL. Furthermore, IL-7 maintains the stemness and self-renewal capacity of lymphoid progenitor cells [223]. Dysregulated IL-7 signaling may promote the expansion of leukemic stem cells with increased self-renewal capacity, facilitating the propagation of leukemic clones and disease progression in ALL. Lastly, IL-7 influences interactions between lymphoid progenitor cells and the bone marrow microenvironment, regulating cell adhesion, migration, and homing [224]. Dysregulated IL-7 signaling may disrupt these interactions, leading to changes in the bone marrow niche that support leukemic cell survival, proliferation, and chemoresistance. These alterations contribute to disease progression in ALL by creating a microenvironment conducive to leukemic cell survival and expansion [225].

##### Down Syndrome based mutations (GATA1, CRLF2, JAK2, RAS) in ALL Leukemogenesis and in increasing ALL Risk

GATA1 mutations in DS-ALL could potentially dysregulate IL-7 expression either directly or indirectly by altering transcriptional regulation. GATA1, a transcription factor, has the ability to modulate the activity of regulatory elements controlling IL-7 gene expression [226, 227]. This dysregulation of IL-7 expression due to GATA1 mutations, combined with potential dosage effects in DS, may disrupt normal lymphoid development. Altered IL-7 levels resulting from GATA1 mutations can potentially promote Lymphoid Progenitor Cell (LPC) survival, proliferation, and impaired differentiation, thereby contributing to the pathogenesis of ALL [228]. Similarly, dysregulated CRLF2 signaling in DS-ALL can indirectly influence IL-7 expression by modulating signaling pathways involved in its regulation. Changes in IL-7 expression driven by dysregulated CRLF2 signaling, in the context of DS, have the potential to disrupt normal lymphoid development [229]. Aberrant IL-7 levels may consequently promote leukemic cell survival, proliferation, and impaired differentiation, thereby contributing to the development of ALL. Moreover, JAK2 mutations in DS-ALL activate downstream signaling pathways, including JAK/STAT, which can directly modulate IL-7 expression. Dysregulated JAK2 signaling may also indirectly affect IL-7 expression through regulatory factors. This dysregulation of IL-7 expression driven by JAK2 mutations, in the context of DS, may interfere with normal lymphoid development, promoting aberrant LPC survival, proliferation, and impaired differentiation, thereby contributing to ALL pathogenesis [230]. Additionally, mutations in the RAS pathway, such as activating mutations in RAS genes, may impact IL-7 expression through downstream signaling cascades involved in its regulation. Dysregulated RAS signaling can potentially modulate IL-7 levels indirectly through its effectors. Consequently, dysregulated IL-7 expression resulting from RAS pathway mutations, in the context of DS, may disrupt normal lymphoid development. Altered IL-7 levels may then promote leukemic cell survival, proliferation, and impaired differentiation, thereby contributing to the development of ALL [231, 232].

#### 13. Stem Cell Factor Signaling

##### Impact of Dysregulations in Disrupting the Cell Fate of Lymphoid Progenitor Cells

Stem Cell Factor (SCF), also referred to as Kit ligand, plays a crucial role in the development and maintenance of hematopoietic stem cells (HSCs) and progenitor cells, including Lymphoid Progenitor Cells (LPCs) [233]. Dysregulation of SCF signaling has the potential to disrupt normal lymphoid development and potentially contribute to the development of Acute Lymphoblastic Leukemia (ALL). There are several possible ways dysregulations in SCF may disrupt the cell fate of LPCs towards ALL [234]. SCF normally promotes LPC survival and proliferation by activating signaling pathways such as PI3K/AKT and MAPK. However, aberrant SCF signaling, such as overproduction of SCF or dysregulated receptor expression, can lead to excessive LPC survival and proliferation. This dysregulation creates a pool of pre-leukemic cells with increased proliferation potential, a hallmark of ALL. Additionally, SCF is involved in maintaining the stemness and self-renewal capacity of HSCs and LPCs [235]. Dysregulated SCF signaling may enhance LPC self-renewal capacity, thereby promoting the expansion of leukemic stem cells with increased potential for long-term survival and propagation of leukemic clones in ALL. Furthermore, SCF influences LPC differentiation by regulating the expression of lineage-specific transcription factors. Dysregulated SCF signaling may disrupt normal LPC differentiation, resulting in the accumulation of undifferentiated or partially differentiated cells with blocked differentiation pathways. This disruption contributes to the development of ALL characterized by the presence of immature lymphoid blasts [236]. Moreover, SCF mediates interactions between LPCs and the bone marrow microenvironment, including stromal cells and extracellular matrix components. Dysregulated SCF signaling can alter these interactions, leading to changes in LPC adhesion, migration, and responsiveness to microenvironmental cues. These changes create a permissive niche for the expansion and survival of leukemic cells in ALL. Finally, SCF signaling influences LPC survival by modulating apoptotic pathways. Dysregulated SCF signaling may disrupt apoptotic pathways, leading to increased survival of pre-leukemic LPCs and resistance to cell death signals. This dysregulation contributes to leukemic transformation and maintenance in ALL [237].

##### Damage leading to ALL Leukemogenesis

SCF, or Stem Cell Factor, is crucial in activating signaling pathways such as PI3K/AKT and MAPK, thereby promoting cell proliferation and survival in Lymphoid Progenitor Cells (LPCs). However, dysregulated SCF signaling, stemming from overexpression of SCF or aberrant receptor expression, can trigger constitutive activation of these pathways, fostering uncontrolled cell proliferation characteristic of ALL [238]. Additionally, SCF is crucial for maintaining the stemness and self-renewal capacity of LPCs. When SCF signaling is dysregulated, it may enhance LPC self-renewal capacity, fostering the expansion of leukemic stem cells with an elevated potential for long-term survival and propagation of leukemic clones in ALL [239]. Furthermore, SCF influences LPC differentiation by regulating the expression of lineage-specific transcription factors (TFs). Dysregulated SCF signaling may disrupt this process, resulting in the accumulation of undifferentiated or partially differentiated cells, a hallmark of ALL. Moreover, SCF plays a key role in mediating interactions between LPCs and the bone marrow microenvironment. Dysregulated SCF signaling can disrupt these interactions, causing changes in LPC adhesion, migration, and responsiveness to microenvironmental cues [240, 241]. These alterations can foster leukemic cell survival and proliferation in the context of ALL. Finally, SCF signaling influences LPC survival by modulating apoptotic pathways. Dysregulated SCF signaling may disrupt these pathways, leading to increased survival of pre-leukemic LPCs and resistance to cell death signals. This phenomenon contributes to leukemic transformation and maintenance in ALL [242, 243].

##### Down Syndrome based mutations (GATA1, CRLF2, JAK2, RAS) in ALL Leukemogenesis and in increasing ALL Risk

GATA1 mutations in DS-ALL have the potential to directly or indirectly dysregulate SCF expression by altering transcriptional regulation. GATA1, as a regulatory factor, can modulate the activity of elements controlling SCF gene expression. Consequently, dysregulated SCF expression due to GATA1 mutations, coupled with potential dosage effects in DS, may disturb normal lymphoid development [244]. This alteration in SCF levels can contribute to ALL pathogenesis by promoting Lymphoid Progenitor Cell (LPC) survival, proliferation, and impaired differentiation. Similarly, dysregulated CRLF2 signaling in DS-ALL can indirectly influence SCF expression by modulating signaling pathways involved in its regulation. Altered CRLF2 signaling may impact SCF levels through downstream effectors such as the JAK/STAT pathways [245]. Changes in SCF expression driven by dysregulated CRLF2 signaling, in the context of DS, can disrupt normal lymphoid development. Consequently, aberrant SCF levels may contribute to ALL development by promoting leukemic cell survival, proliferation, and impaired differentiation. Moreover, JAK2 mutations in DS-ALL activate downstream signaling pathways, including JAK/STAT, which can modulate SCF expression. Dysregulated JAK2 signaling may directly influence SCF transcription or indirectly affect its expression through regulatory factors [246]. As a result, dysregulated SCF expression driven by JAK2 mutations, in the context of DS, may disrupt normal lymphoid development. Altered SCF levels can then promote aberrant LPC survival, proliferation, and impaired differentiation, contributing to ALL pathogenesis. Furthermore, mutations in the RAS pathway, such as activating mutations in RAS genes, may impact SCF expression through downstream signaling cascades involved in its regulation [247, 248]. Dysregulated RAS signaling can modulate SCF levels indirectly through its effectors. Consequently, dysregulated SCF expression resulting from RAS pathway mutations, in the context of DS, can disrupt normal lymphoid development. Altered SCF levels may then promote leukemic cell survival, proliferation, and impaired differentiation, thereby contributing to ALL development [249, 250].

#### 14. Epigenetic Modifiers

##### Impact of Dysregulations in Disrupting the Cell Fate of Lymphoid Progenitor Cells

Epigenetic modifiers are essential in regulating gene expression patterns and chromatin structure during cellular differentiation and development, including in Lymphoid Progenitor Cells (LPCs) [251]. Dysregulation of these modifiers can disrupt normal lymphoid development and contribute to the development of Acute Lymphoblastic Leukemia (ALL) [252]. Dysregulations in epigenetic modifiers can disrupt the cell fate of LPCs towards ALL in several ways. Epigenetic modifiers such as DNA methyltransferases (DNMTs), play a crucial role in regulating DNA methylation patterns, which are essential for gene expression regulation and cellular identity [253]. Dysregulated DNMT activity can result in aberrant DNA methylation patterns, including hypermethylation of tumor suppressor genes or hypomethylation of oncogenes, consequently altering gene expression profiles and contributing to ALL development. Secondly, epigenetic modifiers, including histone acetyltransferases (HATs) and histone deacetylases (HDACs), govern histone modifications, such as acetylation and methylation, which influence chromatin structure and gene accessibility. Imbalances in histone modifications, such as decreased histone acetylation or increased histone methylation, can lead to altered chromatin accessibility and aberrant gene expression patterns, thus promoting leukemogenesis in ALL [254]. Thirdly, epigenetic modifiers, such as ATP-dependent chromatin remodeling complexes, regulate chromatin structure and accessibility, facilitating transcription factor binding and gene expression regulation. Dysregulated chromatin remodeling activities can result in aberrant chromatin structures and impaired transcriptional regulation, disrupting normal LPC differentiation and contributing to ALL pathogenesis [255]. Moreover, epigenetic modifiers also regulate microRNA expression, which modulates gene expression post-transcriptionally. Dysregulated microRNA expression profiles can alter the expression of key regulators of hematopoiesis and leukemogenesis, thereby promoting the expansion of leukemic cells and inhibiting differentiation in ALL [256]. Additionally, epigenetic modifiers influence the expression and function of long non-coding RNAs (lncRNAs), which play diverse roles in gene regulation, chromatin organization, and cellular processes. Dysregulated lncRNA expression can impact gene expression programs and cellular processes critical for LPC differentiation and leukemogenesis, thereby contributing to ALL development [257, 258].

##### Damage leading to ALL Leukemogenesis

Epigenetic modifiers play crucial roles in regulating various aspects of gene expression and chromatin structure during hematopoietic differentiation, including in Lymphoid Progenitor Cells (LPCs) [259]. Dysregulation of these modifiers can significantly impact normal lymphoid development and contribute to the pathogenesis of Acute Lymphoblastic Leukemia (ALL). Dysregulations in epigenetic modifiers can disrupt LPC fate towards ALL through several mechanisms. Epigenetic modifiers regulate the expression of transcription factors (TFs) crucial for hematopoietic differentiation and lineage commitment [260]. Dysregulated expression of TFs such as E2A (TCF3), EBF1, and Ikaros, mediated by epigenetic modifiers, can hinder normal lymphoid differentiation and promote the expansion of leukemic progenitors in ALL. Secondly, epigenetic modifiers influence signaling pathways essential for LPC development and function, including Notch signaling and cytokine receptor pathways. Dysregulated epigenetic modifiers can lead to aberrant activation of signaling pathways like Notch, Wnt, and JAK/STAT, promoting leukemic cell expansion and inhibiting differentiation, thus contributing to ALL pathogenesis. Additionally, epigenetic modifiers regulate DNA methylation patterns, which affect gene expression by modulating chromatin accessibility and TF binding [261, 262]. Dysregulated DNA methylation patterns, caused by epigenetic modifiers, can result in the silencing of tumor suppressor genes or activation of oncogenes critical for leukemic transformation and progression in ALL. Furthermore, epigenetic modifiers control histone modifications, such as acetylation and methylation, which regulate chromatin structure and gene expression [263]. Dysregulated histone modifications, including imbalances in acetylation and methylation, can alter chromatin accessibility and gene expression patterns associated with ALL leukemogenesis [264]. Epigenetic modifiers regulate the expression of non-coding RNAs, including microRNAs and long non-coding RNAs, which modulate gene expression post-transcriptionally. Dysregulated expression of non-coding RNAs involved in regulating key signaling pathways and TFs can contribute to ALL development and progression by influencing LPC fate determination [265].

##### Down Syndrome based mutations (GATA1, CRLF2, JAK2, RAS) in ALL Leukemogenesis and in increasing ALL Risk

GATA1 mutations in DS-ALL may interfere with the normal regulatory functions of Epigenetic Modifiers, such as DNA methyltransferases (DNMTs) and histone modifiers, potentially by altering their expression or activity levels [266]. This disruption could contribute to aberrant DNA methylation patterns, histone modifications, and chromatin remodeling, ultimately influencing gene expression profiles critical for Lymphoid Progenitor Cell (LPC) differentiation and the development of Acute Lymphoblastic Leukemia (ALL) [267]. Similarly, dysregulated CRLF2 signaling in DS-ALL might indirectly affect Epigenetic Modifiers by modulating downstream signaling pathways involved in their regulation. Perturbations in cytokine receptor signaling cascades could lead to altered expression or activity of Epigenetic Modifiers, resulting in epigenetic alterations that impact LPC differentiation and the leukemic transformation process, thereby contributing to disease pathogenesis [268, 269]. JAK2 mutations in DS-ALL can dysregulate Epigenetic Modifiers through the aberrant activation of downstream signaling pathways, particularly the JAK/STAT pathway, which can modulate their expression or activity levels. Consequently, dysregulated Epigenetic Modifiers may induce altered epigenetic landscapes, including changes in DNA methylation, histone modifications, and chromatin accessibility, thereby promoting leukemogenesis in ALL [270, 271]. Moreover, mutations in the RAS pathway in DS-ALL can influence Epigenetic Modifiers by modulating downstream signaling cascades that intersect with epigenetic regulatory mechanisms. This dysregulation may lead to epigenetic alterations favoring leukemic transformation, including aberrant gene expression patterns and chromatin remodeling events associated with the pathogenesis of ALL [272, 273].

## Discussion

### Step-by-Step Origin of Acute Lymphoblastic Leukemia (ALL)

#### Genetic Predisposition and Initial Mutations

Individuals, particularly those with Down Syndrome, possess a genetic predisposition to ALL due to chromosomal abnormalities, such as trisomy 21. Initial mutations occur in key genes like GATA1, CRLF2, JAK2, and RAS. These mutations can occur in hematopoietic stem cells or early lymphoid progenitor cells (LPCs), setting the stage for leukemogenesis.

#### Impact on Key Transcription Factors

Mutations in GATA1, CRLF2, JAK2, and RAS lead to dysregulation of critical transcription factors like Ikaros, PU.1, E2A (TCF3), EBF1, and Notch1. These transcription factors are essential for normal LPC proliferation and differentiation.

#### Disruption of Signaling Pathways

Aberrations in signaling pathways involving IL-7, SCF, FLT3 ligand, and the Notch pathway further contribute to the disrupted development of LPCs. These pathways are crucial for maintaining the balance between LPC proliferation and differentiation.

#### Epigenetic Modifications

Dysregulated expression and function of epigenetic modifiers such as DNMTs, HDACs, and HATs alter the chromatin landscape, leading to inappropriate gene expression patterns that promote leukemic transformation.

#### Proliferation Dysregulations

Enhanced expression of surface markers CD34 and CD38, along with increased activity of IL-7, SCF, and FLT3 ligand, drive the excessive proliferation of LPCs. This uncontrolled proliferation is a hallmark of ALL development.

#### Impaired Differentiation

Dysregulated transcription factors (Ikaros, PU.1, E2A, EBF1, Notch1) and altered signaling pathways (IL-7, Notch) hinder the proper differentiation of LPCs into mature lymphoid cells. Abnormal expression of differentiation markers such as CD10, CD7, and CD45RA disrupts normal lymphoid lineage commitment, leading to an accumulation of immature, non-functional lymphoblasts.

#### Clonal Expansion

The combination of enhanced proliferation and impaired differentiation results in the clonal expansion of these aberrant LPCs. This clonal population eventually dominates the bone marrow, crowding out normal hematopoietic cells.

#### Full-Blown Leukemogenesis

The accumulation of genetic, epigenetic, and signaling pathway dysregulations culminates in the full transformation of LPCs into leukemic blasts. These leukemic cells exhibit uncontrolled growth and impaired differentiation, characteristic of ALL. The leukemic blasts proliferate uncontrollably, leading to the clinical manifestations of ALL, including anemia, thrombocytopenia, and immune suppression due to bone marrow failure.

#### Clinical Presentation and Diagnosis

Patients present with symptoms such as fatigue, frequent infections, easy bruising, and bleeding. Diagnosis is confirmed through bone marrow biopsy, blood tests, and genetic profiling to identify specific mutations and dysregulations.

#### Implications for Treatment

Understanding the step-by-step origin of ALL provides a foundation for developing targeted therapies that address specific genetic and molecular abnormalities. Treatment strategies may include targeted inhibitors for mutated genes, epigenetic therapies, and interventions that restore normal signaling pathways and transcription factor functions.

### Proliferation vs. Differentiation in ALL Leukemogenesis

#### Proliferation Component

Dysregulations in Acute Lymphoblastic Leukemia (ALL) leukemogenesis often manifest as enhanced proliferation signals and reduced differentiation cues within Lymphoid Progenitor Cells (LPCs). This dysregulation involves various factors contributing to uncontrolled LPC proliferation. Factors such as FLT3 Ligand, IL-7, and Stem Cell Factor (SCF), known for their role in promoting proliferation, exhibit increased expression or activity in this context. Additionally, dysregulated epigenetic modifiers like DNA methyltransferases (DNMTs), histone deacetylases (HDACs), and histone acetyltransferases (HATs) can further drive uncontrolled proliferation by altering chromatin structure and gene expression patterns. Moreover, dysregulation of critical transcription factors such as Ikaros and PU.1, which play significant roles in lymphoid lineage commitment and differentiation, can contribute to enhanced LPC proliferation. Dysfunctional or aberrantly expressed transcription factors may disrupt the normal regulatory networks governing LPC proliferation and differentiation processes. This disruption exacerbates the proliferation component of leukemogenesis. Furthermore, alterations in surface markers commonly associated with LPCs, such as CD34 and CD38, are observed in ALL. Changes in the expression patterns of these markers reflect the dysregulated state of LPCs, supporting enhanced proliferation and impaired differentiation. These altered surface marker profiles serve as additional indicators of the dysregulated proliferation component in ALL leukemogenesis.

#### Differentiation Component

In Acute Lymphoblastic Leukemia (ALL), dysregulations often lead to impaired differentiation of Lymphoid Progenitor Cells (LPCs), resulting in the accumulation of undifferentiated or partially differentiated progenitor cells. This impairment involves various factors that disrupt the normal progression of LPCs towards mature lymphoid lineages. The Notch signaling pathway, known for its role in promoting differentiation, can be dysregulated in ALL, hindering LPC differentiation. Similarly, transcription factors like E2A (TCF3), EBF1, and Notch1, which are crucial for orchestrating lymphoid differentiation, may exhibit aberrant expression or function, further contributing to differentiation defects. Moreover, alterations in the expression of differentiation-related surface markers, such as CD10, CD7, and CD45RA, are observed in ALL. These changes in surface marker profiles often signify a blockage or disruption in LPC differentiation pathways, reflecting the impaired differentiation component of ALL leukemogenesis. Additionally, dysregulated epigenetic modifiers, including DNA methyltransferases (DNMTs), histone deacetylases (HDACs), and histone acetyltransferases (HATs), play a role in regulating gene expression during differentiation. In ALL, dysregulation of these epigenetic modifiers may lead to the silencing of genes essential for lymphoid differentiation, exacerbating differentiation defects observed in the disease.

#### Down Syndrome Interplay

Mutations in GATA1, CRLF2, JAK2, and RAS can have diverse effects on the balance between proliferation and differentiation in Lymphoid Progenitor Cells (LPCs), depending on the specific mutation and its downstream consequences. Starting with GATA1 mutations, these alterations can lead to increased proliferation signals in LPCs by dysregulating downstream targets involved in LPC proliferation. This dysregulation may manifest through altered expression of surface markers like CD34 and CD38 and signaling molecules such as IL-7 and SCF. On the other hand, GATA1 mutations can impair LPC differentiation by disrupting the expression or function of transcription factors critical for lymphoid lineage commitment, like PU.1 and Notch1, and by influencing epigenetic modifiers like DNMTs and HDACs. Moving to CRLF2 alterations, changes in CRLF2 signaling, whether through overexpression or activating mutations, can promote LPC proliferation by enhancing signaling through cytokine receptors like FLT3 and IL-7R. Similarly, dysregulated CRLF2 signaling may impact LPC differentiation pathways, potentially by modulating the expression or activity of transcription factors like E2A (TCF3) and epigenetic modifiers involved in regulating lymphoid differentiation. JAK2 mutations, on the other hand, can hyperactivate downstream signaling pathways, such as the JAK/STAT pathway, leading to increased LPC proliferation by upregulating surface markers like CD34 and CD38 and signaling molecules like IL-7 and FLT3 Ligand. Meanwhile, dysregulated JAK2 signaling may interfere with LPC differentiation programs by affecting the expression or function of transcription factors like PU.1 and Notch1, as well as epigenetic modifiers that regulate lymphoid lineage specification. Lastly, mutations in the RAS pathway, including activating mutations in RAS genes, can drive LPC proliferation by promoting aberrant signaling through downstream effectors like RAF and MEK. Dysregulated RAS signaling may perturb LPC differentiation pathways by influencing the expression or activity of transcription factors like EBF1 and Notch1, as well as epigenetic modifiers involved in regulating lymphoid lineage commitment. The mutations in GATA1, CRLF2, JAK2, and RAS often disrupt the balance between LPC proliferation and differentiation, contributing to the pathogenesis of Acute Lymphoblastic Leukemia (ALL). These mutations typically exert their effects through dysregulation of key components of the LPC combinatorial code, including transcription factors, surface markers, signaling molecules, and epigenetic modifiers.

### Landscape of ALL based on the cell-type specific programming

#### Implications of this study

The implications of this study are far-reaching and provide valuable insights into the pathogenesis of Acute Lymphoblastic Leukemia (ALL) and potential therapeutic strategies. By elucidating the critical mechanisms underlying dysregulations in Lymphoid Progenitor Cells (LPCs), it has identified several key implications. The findings pinpoint specific transcription factors, surface markers, signaling molecules, and epigenetic modifiers that are dysregulated in ALL. These molecular targets represent potential candidates for transformative therapeutic interventions aimed at restoring normal LPC function and inhibiting leukemic transformation. Understanding the genetic architecture of LPCs in ALL allows for the development of targeted therapies tailored to specific molecular abnormalities. Targeted agents, such as small molecule inhibitors or monoclonal antibodies, can selectively block aberrant signaling pathways or epigenetic modifications, offering more effective and less toxic treatment options for patients with ALL. This study highlights the heterogeneity of ALL and the importance of personalized medicine approaches. By characterizing the genetic profiles of individual patients, clinicians can tailor treatment strategies based on the unique molecular abnormalities present in their disease, leading to improved outcomes and reduced side effects. By dissecting the molecular pathways involved in ALL development, we gain insights into disease progression and mechanisms of treatment resistance. This can inform the development of combination therapies targeting multiple dysregulated pathways simultaneously, thereby overcoming resistance and improving treatment efficacy. The molecular signatures identified in this study may have prognostic implications, allowing for more accurate risk stratification of patients with ALL. By integrating genetic, epigenetic, and clinical data, clinicians can better predict disease outcomes and tailor treatment intensity accordingly, optimizing patient care and long-term survival. This study provides crucial understanding of the genetic underpinnings of ALL and their implications for disease pathogenesis and treatment. The insights gained from this study have the potential to revolutionize the management of ALL, leading to more effective therapies, improved patient outcomes, and ultimately, a better quality of life for individuals affected by this devastating disease.

#### Clinical Implications

The findings of this research hold several important clinical implications for the diagnosis, treatment, and management of Acute Lymphoblastic Leukemia (ALL). Identification of specific genetic abnormalities, such as mutations in transcription factors (e.g., Ikaros, PU.1) and surface markers (e.g., CD34, CD38), can serve as diagnostic biomarkers for ALL. Clinicians can use these biomarkers to aid in the accurate and early diagnosis of the disease. Certain genetic alterations identified in this research may serve as prognostic indicators, helping clinicians predict disease outcomes and tailor treatment plans accordingly. For example, the presence of certain mutations or dysregulations may indicate a higher risk of relapse or resistance to standard therapies. Understanding leukemogenesis involved in ALL development allows for the development of targeted therapies that specifically inhibit dysregulated signaling pathways or epigenetic modifications. Targeted therapies can improve treatment efficacy while minimizing toxicity compared to traditional chemotherapy approaches. By analyzing the genetic profile of individual patients, clinicians can personalize treatment approaches based on the specific molecular abnormalities present in their disease. This personalized approach maximizes treatment efficacy while minimizing unnecessary exposure to potentially toxic therapies. Molecular profiling of ALL can be used to monitor treatment response and detect minimal residual disease (MRD) during and after therapy. Clinicians can use biomarkers associated with specific genetic abnormalities to assess treatment effectiveness and adjust therapy as needed to optimize patient outcomes. The identification of key genetic targets and pathways in ALL opens up opportunities for clinical trials and further research into new transformative treatment approaches. Investigational therapies targeting specific genetic abnormalities identified in this research may offer promising avenues for improving outcomes in patients with ALL. Translating the findings of this research into clinical practice has the potential to revolutionize the diagnosis, treatment, and management of ALL, ultimately leading to better outcomes and quality of life for patients affected by this disease.

#### Key Findings of the Study

This study uncovered significant findings regarding the interplay between genetic dysregulations and proliferation in Acute Lymphoblastic Leukemia (ALL). Mutations in GATA1, CRLF2, JAK2, and RAS genes were found to significantly affect the expression of key transcription factors like Ikaros and PU.1, as well as surface markers such as CD34 and CD38. This discovery highlights a direct mechanistic link between these mutations and the upregulation of proliferative signals in lymphoid progenitor cells (LPCs). Additionally, the study provided new evidence that these mutations not only enhance proliferative signals but also disrupt differentiation pathways. Dysregulation of transcription factors such as E2A, EBF1, and Notch1, along with surface markers like CD10, CD7, and CD45RA, impedes normal LPC differentiation, contributing to leukemic transformation. This dual impact on both proliferation and differentiation was previously underappreciated. This study also revealed that mutations in Down Syndrome patients alter the activity of epigenetic modifiers such as DNMTs, HDACs, and HATs, resulting in aberrant chromatin remodeling and gene expression patterns that favor leukemogenesis. The specific epigenetic pathways influenced by these mutations were not fully understood before this study. Furthermore, transformative insights were gained into how IL-7, SCF, and FLT3 ligand signaling pathways are affected by genetic mutations in Down Syndrome. The study demonstrated that these signaling molecules are upregulated, leading to enhanced survival and proliferation of LPCs, thereby promoting leukemogenesis. The precise contribution of these pathways in the context of Down Syndrome was previously unclear. This research provides an integrated model showing how genetic, epigenetic, and signaling pathway alterations collectively drive the leukemic transformation of LPCs. It emphasizes the importance of a multidimensional approach to understanding leukemogenesis, incorporating genetic mutations, signaling dysregulations, and epigenetic changes. Specifically, the study delineated how Down Syndrome-associated mutations (GATA1, CRLF2, JAK2, and RAS) uniquely contribute to ALL pathogenesis. This includes new insights into the differential expression of genes and pathways in Down Syndrome compared to other genetic contexts, shedding light on why Down Syndrome patients have a higher predisposition to ALL. These findings collectively advance the understanding of ALL leukemogenesis, particularly in the context of Down Syndrome, and open new avenues for targeted therapeutic interventions.

#### Acute Lymphoblastic Leukemia and Cell-Type Specific Programming of Lymphoid Progenitor Cells

The stem cells that give rise to ALL are primarily early lymphoid progenitors derived from HSCs. Acute Lymphoblastic Leukemia (ALL) is a malignancy of the bone marrow in which early lymphoid progenitor cells undergo malignant transformation. These transformed cells proliferate uncontrollably, leading to the accumulation of immature lymphoblasts. HSCs give rise to two major progenitor lineages, myeloid progenitors and lymphoid progenitors. Common Lymphoid Progenitors (CLPs) are derived from HSCs and are committed to the lymphoid lineage. CLPs have the potential to differentiate into B-cell progenitors, T-cell progenitors, and natural killer (NK) cell progenitors. ALL originates from cells committed to the lymphoid lineage, specifically early B-cell and T-cell progenitors. The pathogenesis involves a complex interplay of genetic and epigenetic changes that drive malignant transformation, proliferation, and survival. Understanding these processes is crucial for developing targeted therapies and improving patient outcomes. Research continues to focus on identifying leukemic stem cells and their unique properties to devise strategies that prevent relapse and achieve lasting remissions. Both B-ALL and T-ALL arise due to genetic and epigenetic alterations in these progenitor cells, leading to uncontrolled proliferation and accumulation of lymphoblasts. Understanding this lineage commitment is crucial for comprehending the origins and development of diseases like ALL, which result from malignant transformations within these progenitor populations. Common Lymphoid Progenitors (CLPs) are also known as Lymphoid Progenitor Cells.

The cell-type specific programming of Lymphoid Progenitor Cells, or Common Lymphoid Progenitors (CLPs), refers to the specific combination of transcription factors, surface markers, and signaling molecules that define their identity and guide their differentiation into various lymphoid cell types. Key transcription factors involved in this programming include Ikaros, PU.1, E2A (TCF3), EBF1, and Notch1. The surface markers that characterize these cells are CD34, CD38, CD10, CD7, CD45RA, and FLT3 (CD135). Important signaling molecules such as Interleukin-7 (IL-7) and Stem Cell Factor (SCF) play crucial roles in their development. Additionally, epigenetic modifiers like DNA Methyltransferases (DNMTs) and histone modifiers (e.g., HDACs, HATs) are essential for regulating the gene expression patterns necessary for their differentiation. Understanding this critical programming is crucial for comprehending how CLPs maintain their multipotency and how they differentiate into specific lymphoid cell types, as well as for devising targeted therapies in diseases such as acute lymphoblastic leukemia (ALL). The combinatorial code of Lymphoid Progenitor Cells (CLPs) involves factors related to their roles in proliferation and differentiation. Proliferation-related factors are primarily involved in promoting the survival, expansion, and proliferation of CLPs. Transcription factors such as Ikaros and PU.1 play roles in regulating genes that control cell cycle and proliferation, with PU.1 also functioning in early progenitor proliferation. Surface markers like CD34 and CD38 are associated with proliferating hematopoietic progenitors and activated, proliferating progenitor cells, respectively. Signaling molecules such as Interleukin-7 (IL-7), which activates the JAK/STAT pathway, and Stem Cell Factor (SCF), which binds to the c-Kit receptor, promote proliferation and survival of CLPs. The FLT3 Ligand stimulates the FLT3 receptor to support the expansion of CLPs by activating the FLT3 signaling pathway. Epigenetic modifiers, including DNA Methyltransferases (DNMTs) and histone modifiers (e.g., HDACs, HATs), regulate the methylation status of DNA and modify chromatin structure to control the accessibility of proliferation-related genes. Differentiation-related factors guide the differentiation of CLPs into specific lymphoid lineages, such as B-cells, T-cells, and NK cells. Transcription factors like Ikaros, which is essential for lymphoid differentiation, and PU.1, which directs the differentiation of lymphoid progenitors, particularly B-cell and T-cell lineages, play critical roles. E2A (TCF3) and EBF1 (Early B-cell Factor 1) are crucial for early B-cell lineage commitment and differentiation, with EBF1 working with E2A to promote B-cell differentiation. Notch1 is vital for T-cell lineage commitment and differentiation by activating T-cell-specific genes and repressing B-cell fate. Surface markers such as CD10 (CALLA), involved in B-cell differentiation, CD7, a marker of early T-cell progenitors, and CD45RA, involved in the differentiation of CLPs, are important indicators of lineage commitment. Signaling molecules like IL-7, which also promotes T-cell differentiation, and the Notch signaling pathway, which directs T-cell lineage differentiation and inhibits B-cell development in thymic progenitors, are critical for guiding CLP differentiation. Epigenetic modifiers, including DNMTs and histone modifiers, control chromatin accessibility for genes involved in differentiation processes and regulate differentiation-specific gene expression.

#### Impact of Down Syndrome

Down Syndrome significantly affects the combinatorial code of Lymphoid Progenitor Cells by altering the expression and function of transcription factors, surface markers, signaling molecules, and epigenetic modifiers. These alterations lead to disrupted proliferation and differentiation of CLPs, contributing to the increased risk of developing acute lymphoblastic leukemia (ALL) in individuals with DS. Understanding these changes is crucial for developing targeted therapies and improving outcomes for DS-associated leukemias. Down Syndrome (DS), caused by trisomy 21, is associated with various hematological abnormalities, including an increased risk of leukemia, particularly acute lymphoblastic leukemia (ALL). The combinatorial code of Lymphoid Progenitor Cells (CLPs) is significantly affected in individuals with Down Syndrome.

In terms of proliferation roles, transcription factors such as Ikaros (IKZF1) and PU.1 (SPI1) are impacted. DS leads to overexpression of genes on chromosome 21, which can disrupt Ikaros activity, affecting its regulatory roles in proliferation, while PU.1 expression may be indirectly affected due to altered signaling pathways and chromosomal abnormalities, potentially impacting progenitor cell proliferation. Surface markers like CD34 and CD38 are also affected, with overexpression of chromosome 21 genes altering their expression and impacting the proliferation of CLPs. Signaling molecules, including Interleukin-7 (IL-7), Stem Cell Factor (SCF), and FLT3 Ligand, have disrupted signaling pathways in DS, affecting the proliferation and survival of lymphoid progenitors. Epigenetic modifiers such as DNA Methyltransferases (DNMTs) and histone modifiers (HDACs, HATs) experience altered activity, leading to abnormal DNA methylation patterns and dysregulated gene expression, which impact cell proliferation.

For differentiation roles, transcription factors like Ikaros (IKZF1) and PU.1 (SPI1) also play crucial roles. Disruption in Ikaros function affects not only proliferation but also differentiation pathways, leading to improper lymphoid development. Altered PU.1 expression impacts both B-cell and T-cell differentiation, while E2A (TCF3) and EBF1 are essential for B-cell lineage commitment and differentiation and may be compromised in DS. Notch1 is crucial for T-cell differentiation, and its altered signaling in DS can lead to improper T-cell lineage commitment and differentiation. Surface markers such as CD10, CD7, and CD45RA are affected, with changes in their expression impacting early B-cell progenitor differentiation, T-cell progenitor differentiation, and differentiation pathways of CLPs, respectively. Signaling molecules like IL-7 and the Notch signaling pathway are crucial for T-cell differentiation, and their disrupted signaling in DS affects T-cell development. Epigenetic modifiers like DNMTs and histone modifiers influence gene expression involved in differentiation, with aberrant DNA methylation patterns and dysregulated histone modification impacting differentiation pathways.

Down Syndrome (DS), characterized by an extra copy of chromosome 21, affects various cellular processes, including the proliferation and differentiation of Lymphoid Progenitor Cells (CLPs). DS leads to dysregulation of Ikaros, resulting in impaired regulation of genes critical for CLP proliferation, which may contribute to abnormal growth and survival of progenitor cells. While PU.1 is less directly affected by trisomy 21, the overall altered transcriptional environment in DS can impact PU.1 function, indirectly affecting CLP proliferation. CD34 expression levels may be altered due to the overexpression of genes on chromosome 21, affecting the proliferation and maintenance of hematopoietic progenitors. Similarly, CD38 expression can be influenced by the altered genetic environment in DS, impacting progenitor cell proliferation. IL-7 signaling is often disrupted in DS, leading to reduced proliferation and survival of lymphoid progenitors. SCF signaling through its receptor c-Kit may be less efficient or altered, affecting the proliferation of CLPs. FLT3 signaling is crucial for progenitor expansion, and disruptions in this pathway in DS can impair the proliferation capacity of CLPs. Abnormal DNMT activity in DS results in altered DNA methylation patterns, which can disrupt the expression of genes involved in cell proliferation. Dysregulation of histone modifiers in DS affects chromatin structure, leading to aberrant expression of proliferation-related genes. Dysregulation of Ikaros impacts not only proliferation but also the proper differentiation of lymphoid cells, contributing to leukemia development. Altered PU.1 activity in the DS environment affects the differentiation of both B-cell and T-cell lineages. E2A is essential for B-cell differentiation, and its function may be impaired in DS, leading to defective B-cell lineage commitment. EBF1 is critical for B-cell development, and DS can disrupt EBF1 function, impacting B-cell differentiation. Notch1 signaling, crucial for T-cell differentiation, may be impaired in DS, leading to abnormal T-cell development. Changes in CD10 expression can affect early B-cell differentiation in DS. Altered CD7 expression impacts T-cell progenitor differentiation. Changes in CD45RA expression can affect the differentiation pathways of CLPs. IL-7’s role in T-cell differentiation is critical, and its signaling disruption in DS impacts T-cell development. Altered Notch signaling in DS can lead to improper differentiation of T-cells, contributing to the skewed lymphoid cell populations seen in DS. Epigenetic changes in DS affect the expression of genes necessary for lymphoid differentiation. Dysregulated histone modification patterns can lead to abnormal gene expression profiles affecting differentiation. DS generally impairs proliferation pathways, leading to abnormal growth and survival of lymphoid progenitor cells. It significantly affects differentiation processes, leading to improper lineage commitment and maturation of lymphoid cells, contributing to the development of leukemias such as ALL. Mutations in genes such as GATA1 and alterations in the CRLF2 gene, along with associated mutations affecting pathways involving either the JAK2 or RAS genes, have been strongly linked to Down Syndrome-Associated Acute Lymphoblastic Leukemia (DS-ALL) leukemogenesis. GATA1, CRLF2, JAK2, and RAS are not typically considered part of the combinatorial code of Lymphoid Progenitor Cells (CLPs). Instead, they represent specific genetic factors and signaling pathways that are implicated in leukemogenesis, including Down Syndrome-Associated Acute Lymphoblastic Leukemia (DS-ALL).

## Conclusions

This study into the origins of ALL investigates the critical roles played by transcription factors, surface markers, signaling molecules, and epigenetic modifiers in maintaining the delicate balance between LPC proliferation and differentiation. Dysregulation of these components, either through genetic mutations or alterations in signaling pathways, can disrupt normal hematopoietic development and predispose LPCs to leukemic transformation. Specifically, mutations in genes such as GATA1, CRLF2, JAK2, and RAS have been implicated in driving aberrant LPC proliferation and impairing differentiation, thereby contributing to the onset and progression of ALL. These mutations often exert their effects by modulating the expression or activity of key regulators of LPC fate, including transcription factors like Ikaros, PU.1, E2A (TCF3), and EBF1, as well as signaling molecules like IL-7 and components of the Notch signaling pathway. Furthermore, the focus on how Down Syndrome (DS) based mutations impact LPCs has provided insights into the unique genetic landscape of DS-ALL and its underlying mechanisms. DS-associated alterations in GATA1, CRLF2, JAK2, and RAS can exacerbate dysregulation of LPCs, further tipping the balance towards leukemic transformation. This study underscores the importance of understanding the genetic architecture of LPCs in elucidating the pathogenesis of ALL. By looking into the mechanisms underlying LPC dysregulation, we can identify new therapeutic targets and develop more effective treatment strategies for ALL, ultimately improving patient outcomes in this devastating disease.

The landscape of Acute Lymphoblastic Leukemia (ALL) development is characterized by dysregulations in key factors governing the proliferation and differentiation of Lymphoid Progenitor Cells (LPCs). Proliferation dysregulations involve the upregulation of transcription factors such as Ikaros and PU.1, along with increased expression of surface markers like CD34 and CD38. Elevated levels of signaling molecules IL-7, SCF, and FLT3 Ligand drive LPC proliferation, while alterations in epigenetic modifiers DNMTs, HDACs, and HATs further contribute to uncontrolled cell division. On the differentiation front, dysregulated transcription factors Ikaros, PU.1, E2A (TCF3), EBF1, and Notch1 disrupt LPC differentiation pathways. Aberrant expression of surface markers CD10, CD7, and CD45RA hinders normal lymphoid lineage commitment. Dysregulated signaling through IL-7 and the Notch pathway further impairs differentiation processes. Epigenetic modifications mediated by DNMTs, HDACs, and HATs also play a role in inhibiting LPC differentiation. These dysregulations in proliferation and differentiation factors create a landscape conducive to ALL development, characterized by uncontrolled LPC proliferation and impaired differentiation, ultimately leading to leukemic transformation. The findings of this research hold several important clinical implications for the diagnosis, treatment, and management of Acute Lymphoblastic Leukemia (ALL).

## Abbreviations

NF-κB: Nuclear Factor-kappa B
ALL: Acute Lymphoblastic Leukemia
LPCs: Lymphoid Progenitor Cells
PU.1: Purine-rich Box-1
E2A (TCF3): Transcription Factor 3
EBF1: Early B-cell Factor 1
SCF: Stem Cell Factor
FLT3: Fms-like Tyrosine Kinase 3
IL-7: Interleukin-7
Ikaros: IKAROS family zinc finger 1
CD34: Cluster of Differentiation 34
CD38: Cluster of Differentiation 38
CD10: Cluster of Differentiation 10
CD7: Cluster of Differentiation 7
CD45RA: Cluster of Differentiation 45RA
Notch1: Neurogenic locus notch homolog protein 1
CRLF2: Cytokine Receptor-like Factor 2
JAK2: Janus Kinase 2
RAS: Rat Sarcoma Virus Oncogene

## Declarations

## Ethics declarations

**Ethics approval and consent to participate**

Not applicable.

## Consent for publication

Not applicable.

## Data Availability statement

All data generated or analyzed during this study are included in this article.

## Acknowledgements

“Not applicable”.

## Authors’ Information

1. **Ovais Shafi (OS)\*** is the author of the study and was involved in the idea, concept, design, and methodology of the study, literature search and references. He did the writing, editing, and revision of the manuscript. He was involved in drawing the findings, results, conclusions, implications of the study, interpretation of the data and was involved in all aspects of the study. He prepared and wrote discussion, results, conclusions and all areas of the study. OS extracted and analyzed the data. He was involved in critical evaluation, audit of every aspect of the study, data extraction, adherence of the study to relevant PRISMA guidelines, limitations of the study, references, and all others. He was involved in drawing fig 1. The author read and approved the manuscript.

**Fig 1.**
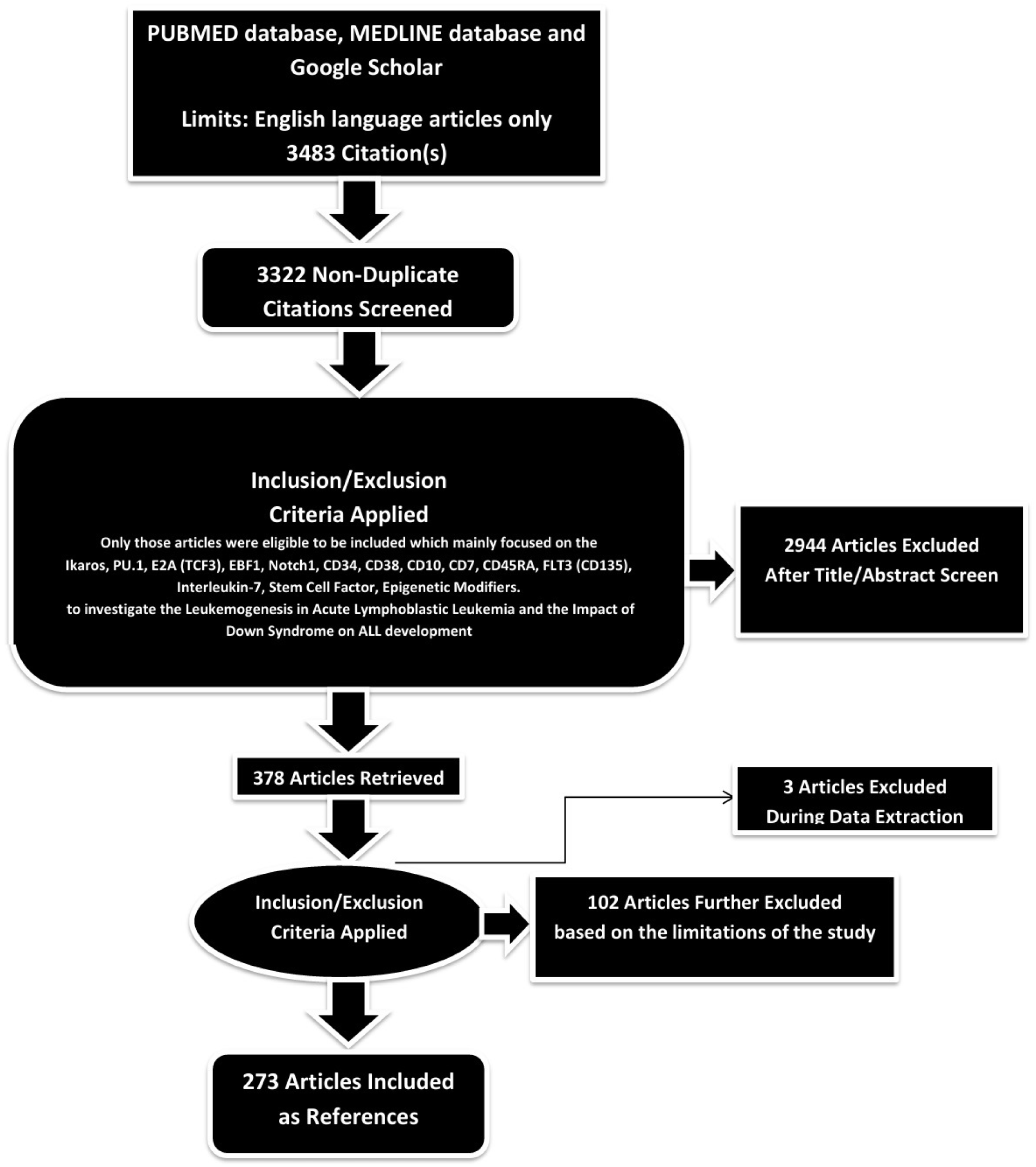
PRISMA FLOW DIAGRAM: This figure represents graphically the flow of citations in the study. He investigated the transcription factors/surface markers/signaling pathways for their roles in relation to this study: Ikaros, PU.1, E2A (TCF3), EBF1, Notch1, CD34, CD38, CD10, CD7, CD45RA, FLT3 (CD135), Interleukin-7, Stem Cell Factor, Epigenetic Modifiers. **Ovais Shafi (OS)\***, MBBS - Sindh Medical College - Dow University of Health Sciences, Karachi, Pakistan. He aspires to become an eminent ‘Physician Scientist’. He is devoted to the research in disease development mechanisms, disease origins and therapeutics. OS is also passionate about multiple research areas including clinical trials, clinical medicine, therapeutics, regenerative medicine, precision medicine including gene therapies, finding disease specific targets for gene therapy, role of disease genomics and epigenetics in diagnosis, management, and therapeutics development. He is dedicated to the field of research and clinical medicine.
2. **Rahimeen Rajpar (RR)** is also the author of the study and contributed to the writing, editing and revision of the study. She also contributed to the results and conclusions of this manuscript along with working on the findings. She contributed to investigating the transcription factors/surface markers/signaling pathways for their roles in relation to this study: Ikaros, PU.1, E2A (TCF3), EBF1, Notch1, CD34, CD38, CD10, CD7, CD45RA, FLT3 (CD135), Interleukin-7, Stem Cell Factor, Epigenetic Modifiers. Rahimeen Rajpar, MD is a dedicated medical professional with a passion for unraveling the mysteries of disease origins and progression. Currently pursuing her residency in internal medicine. RR is committed to advancing the field of medical research. Her goal is to become a leader in the field of Medicine, looking at the medical intricacies from a different lens. Apart from research RR remains dedicated to improving the lives of her patients through comprehensive care and by working towards scientific discoveries. RR is a MBBS graduate from Sindh Medical College – Jinnah Sindh Medical University, Karachi, Pakistan.
3. **Finza Kanwal (FK)** is the co-author of the study. She contributed to the results and conclusions of the study, also contributed to the writing and editing of these sections. She contributed to investigating the transcription factors/surface markers/signaling pathways for their roles in relation to this study: Ikaros, PU.1, E2A (TCF3), EBF1, Notch1, CD34, CD38, CD10, CD7, CD45RA, FLT3 (CD135), Interleukin-7, Stem Cell Factor, Epigenetic Modifiers. Finza Kanwal, MBBS - Sindh Medical College - Dow University of Health Sciences, Karachi, Pakistan. She has been affiliated with Liaquat National Hospital and Medical College, Pakistan. Currently, she is affiliated with The College of Physicians and Surgeons Pakistan. Her goal is to be an Endocrinologist (MRCP).
4. **Muhammad Waqas (MW)** is the co-author of the study. He contributed to the results and conclusions of the study, also contributed to the writing and editing of these sections. He contributed to investigating the transcription factors/surface markers/signaling pathways for their roles in relation to this study: Ikaros, PU.1, E2A (TCF3), EBF1, Notch1, CD34, CD38, CD10, CD7, CD45RA, FLT3 (CD135), Interleukin-7, Stem Cell Factor, Epigenetic Modifiers. Muhammad Waqas, MBBS - Sindh Medical College - Dow University of Health Sciences, Karachi, Pakistan. He is ECFMG Certified. His future goals include residency in Internal Medicine/Neurology, and fellowship in Cardiology/Nephrology/Critical Care.
5. **Osama Jawed Khan (OJK)** is the co-author of the study. He contributed to the results and conclusions of the study, also contributed to the writing and editing of these sections. He contributed to investigating the transcription factors/surface markers/signaling pathways for their roles in relation to this study: Ikaros, PU.1, E2A (TCF3), EBF1, Notch1, CD34, CD38, CD10, CD7, CD45RA, FLT3 (CD135), Interleukin-7, Stem Cell Factor, Epigenetic Modifiers. Osama Jawed Khan, MBBS - Sindh Medical College - Dow University of Health Sciences, Karachi, Pakistan. He has been affiliated with Dr. Ruth K. M. Pfau, Civil Hospital Karachi, Pakistan. Currently, he is affiliated with Royal Institute of Medicine & Surgery (RIMS), Karachi. He intends to pursue fellowship of the Royal Colleges of Surgeons (FRCS).
6. **Raveena (RA)** is the co-author of the study. She contributed to the results and conclusions of the study, also contributed to the writing and editing of these sections. She contributed to investigating the transcription factors/surface markers/signaling pathways for their roles in relation to this study: Ikaros, PU.1, E2A (TCF3), EBF1, Notch1, CD34, CD38, CD10, CD7, CD45RA, FLT3 (CD135), Interleukin-7, Stem Cell Factor, Epigenetic Modifiers. Raveena, MBBS - Sindh Medical College – Jinnah Sindh Medical University, Karachi, Pakistan. She is passionate about research in surgery and disease development mechanisms including neurodegenerative diseases, oncogenesis and others. She is ECFMG Certified. She is passionate about residency in Internal Medicine/Surgery. Her goal is to make significant impact in the field of Research.
7. **Ankash Kumar (AK)** is the co-author of the study. He contributed to the results and conclusions of the study, also contributed to the writing and editing of these sections. He contributed to investigating the transcription factors/surface markers/signaling pathways for their roles in relation to this study: Ikaros, PU.1, E2A (TCF3), EBF1, Notch1, CD34, CD38, CD10, CD7, CD45RA, FLT3 (CD135), Interleukin-7, Stem Cell Factor, Epigenetic Modifiers. Ankash Kumar, MBBS a distinguished graduate of Liaquat University of Medical and Health Sciences, Jamshoro Sindh Pakistan, currently serves in NHS England. With a strong foundation in medical education and clinical practice, Dr. Kumar is committed to advancing his career in internal medicine. His immediate goal is to pursue Residency in Internal Medicine, where he aims to further his skills and be able to contribute towards the field of Medicine.
8. **Abdul Jabbar (AJ)** is the co-author of the study. He contributed to the results and conclusions of the study, also contributed to the writing and editing of these sections. He contributed to investigating the transcription factors/surface markers/signaling pathways for their roles in relation to this study: Ikaros, PU.1, E2A (TCF3), EBF1, Notch1, CD34, CD38, CD10, CD7, CD45RA, FLT3 (CD135), Interleukin-7, Stem Cell Factor, Epigenetic Modifiers. Abdul Jabbar, MBBS - Sindh Medical College - Jinnah Sindh Medical University, Karachi, Pakistan. He is currently a Medical officer in an NGO. He is interested in research in disease development mechanisms. He is ECFMG Certified, his goal is to do Medical residency in Internal Medicine. He aspires to be a specialist in Cardiology.
9. **Manwar Madhwani (MM)** is the co-author of the study. He contributed to the results and conclusions of the study, also contributed to the writing and editing of these sections. He contributed to investigating the transcription factors/surface markers/signaling pathways for their roles in relation to this study: Ikaros, PU.1, E2A (TCF3), EBF1, Notch1, CD34, CD38, CD10, CD7, CD45RA, FLT3 (CD135), Interleukin-7, Stem Cell Factor, Epigenetic Modifiers. Manwar Madhwani, MBBS - Sindh Medical College – Jinnah Sindh Medical University, Karachi, Pakistan. He is currently a medical officer in an NGO. He is interested in research in disease development mechanisms. His goal is to do Medical Residency in Internal Medicine after completing his ECFMG Certification. He aspires to be a specialist in Cardiology/ Pediatrics.

*The work and contributions of everyone have been described in detail, the order is randomized and the numbering is just for referencing purpose. All authors have read and approved the manuscript*.

